# Improved heritability partitioning and enrichment analyses using summary statistics with graphREML

**DOI:** 10.1101/2024.11.04.24316716

**Authors:** Hui Li, Tushar Kamath, Rahul Mazumder, Xihong Lin, Luke O’Connor

## Abstract

Heritability enrichment analysis using data from Genome-Wide Association Studies (GWAS) is often used to understand the functional basis of genetic architecture. Stratified LD score regression (S-LDSC) is a widely used method-of-moments estimator for heritability enrichment, but S-LDSC has low statistical power compared with likelihood-based approaches. We introduce graphREML, a precise and powerful likelihood-based heritability partition and enrichment analysis method. graphREML operates on GWAS summary statistics and linkage disequilibrium graphical models (LDGMs), whose sparsity makes likelihood calculations tractable. We validate our method using extensive simulations and in analyses of a wide range of real traits. On average across traits, graphREML produces enrichment estimates that are concordant with S-LDSC, indicating that both methods are unbiased; however, graphREML identifies 2.5 times more significant trait-annotation enrichments, demonstrating greater power compared to the moment-based S-LDSC approach. graphREML can also more flexibly model the relationship between the annotations of a SNP and its heritability, producing well-calibrated estimates of per-SNP heritability.

## Introduction

Heritability partitioning is a powerful approach to integrate genetic association data with variance functional genomic data^1^, by quantifying the heritability enrichment of a derived annotation. This approach has been used to identify disease-relevant regulatory annotations^2–4^, to prioritize disease-relevant genes and cell types^5, 6^, to investigate the effect of negative selection on genetic architecture^7–10^, and to compare common vs. rare variant architecture^8, 11, 12^.

For common and low-frequency variants, the most widely used heritablity enrichmen method is stratified LD score regression (S-LDSC)^3, 7, 13^. This method is fast, and it operates on publicly available summary association statistics; S-LDSC can also jointly analyze a large number of overlapping annotations. These features distinguish S-LDSC from REML-based methods^14–16^, which require individual level GWAS data and cannot handle overlapping annotations. However, S-LDSC can have much lower statistical power compared with likelihood-based methods, such that many enrichments may go undetected.

This trade-off arises from the difficulty of fully modeling linkage disequilibrium (LD), and in particular, incorporating it into likelihood calculations. S-LDSC relies on “LD scores,” which summarize the LD matrix but result in loss of information^17^. This has motivated various approaches to represent LD parsimoniously, such as shrinkage regularization^18^, banding^19–21^, truncated SVD^22, 23^ or a combination of the latter two^17, 24, 25^. Recently, Nowbandegani and Wohns et al. proposed LD graphical models (LDGMs), which represent LD patterns using extremely sparse matrices derived from genome-wide genealogies^26^. The edge between two adjacent SNPs in the LDGM corresponds to a genealogical relationship between the ancestral haplotypes on which they arise as mutations^27^. LDGMs enable the use of efficient sparse matrix operations to perform likelihood calculations with GWAS data, potentially addressing the challenge of likelihood-based heritability partitioning.

We propose graphREML, a likelihood-based heritability partitioning method that operates on GWAS summary statistics and LDGMs. graphREML improves upon S-LDSC by modeling the full likelihood of the summary statistics, making it more precise and powerful than S-LDSC. Moreover, by directly modeling the likelihood of variant-level summary statistics, graphREML is capable of handling overlapping annotations, unlike the existing REML-based methods which require individual-level data. Because of its higher precision and statistical power, graphREML is particularly advantageous for estimating the heritability enrichment of disease traits which are under-powered using S-LDSC. graphREML is also robust to various forms of model misspecification, e.g., when there is sample mismatch between the GWAS statistics and the LDGM precision matrices.

We validated our method in simulations and in analyses of real traits, comparing the enrichment estimates from our method to those from the S-LDSC. We chose S-LDSC in particular because it is the most widely used method that also operates on summary statistics. One other method that uses summary statistics is SumHer^28^; SumHer fits a different heritability model from S-LDSC, but its inference approach is similar. We also estimated heritability at a per-SNP level, as opposed to at an aggregate level, highlighting the advantages of our approach. Lastly, we note that graphREML can be integrated with other analytical frameworks that utilize enrichment estimates, such as the Abstract Mediation Model (AMM)^29^; this led to a more precise quantification of the degree of mediated heritability by a gene set (*e*.*g*., constrained genes).

## Results

### Overview of graphREML

We propose using a maximum-likelihood approach to estimate partitioned heritability and enrichment. Under the standard assumptions of genetic association modeling, the distribution of the summary association statistics can be derived^17, 30^. Ideally, a maximum-likelihood estimator would be used; however, the likelihood is parameterized by the LD matrix, such that it can be expensive to compute. We exploit the sparsity of the LDGM precision matrices to enable tractable maximization of the GWAS likelihood (**Online Methods**). We employ a second-order optimization method with an approximate Hessian and a trust region algorithm to make the maximization algorithm stable^16^ (**Online Methods**). With these optimizations, the estimation is tractable but still slow, typically requiring 1-3 days for convergence.

A peculiarity of S-LDSC is that for many individual SNPs, its linear heritability model would suggest that their heritability is negative. Because S-LDSC cannot accommodate a non-linear relationship between the heritability of a SNP and its annotations, it cannot enforce non-negativity. In contrast, graphREML uses a non-negative inverse link function to map between the linear combination of the annotations of a SNP and its expected heritability. Let **a** _*j*_ denote the annotation values of SNP *j*. We model the per-SNP heritability of SNP *j* as:

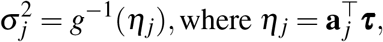

where *τ* is a vector of unknown parameters that encodes the genetic architecture of a trait, and *g*(·) is a non-negative link function^31^. S-LDSC assumes an identity link, *g*(*x*) = *x*. graphREML is guaranteed to produce valid non-negative per-SNP heritability estimates, as long as an appropriate link *g*(·) is applied.

An important feature of S-LDSC is that it can distinguish polygenic effects from confounding due to population stratification and relatedness^13^. graphREML does not model uncorrected population stratification. Instead, it requires the appropriate correction for population stratification either directly at the individual-level (before association testing), or at the summary-statistics level by taking the S-LDSC intercept as an input in order to account for confounding (**Online Methods**).

The marginal heritability enrichment of an annotation may differ from the conditional enrichment. The marginal enrichment can be driven by overlap with other annotations, whereas the conditional enrichment measures the additional enrichment in an annotation after accounting for its overlap with others^3^. A positive conditional enrichment implies that SNPs in that annotation have greater heritability than expected given their other annotations. graphREML (like S-LDSC) estimates both types of enrichment. In this manuscript, we report enrichments estimated under the baselineLD model, which is widely used in conjunction with S-LDSC^7^. This model has been shown to account for frequency-dependent and LD-dependent architecture, which otherwise cause bias when estimating either conditional or marginal heritability enrichments^32^.

We estimate the standard error of enrichment using an approximate jackknife estimator. More specifically, we compute the empirical variance of the leave-one-LD-block-out estimates of the parameters as the jackknife covariance estimator of the conditional enrichment coefficients *τ* (**Online Methods**). The jackknife procedure is computationally efficient, not requiring the model to be re-fitted. For significance testing, we adopt a similar procedure as S-LDSC, applying a Wald test with jackknife standard errors to the difference, rather than the ratio, between the per-SNP heritability in versus out of an annotation. We apply the Delta method to obtain the asymptotic variance of these enrichment test statistics (**Online Methods**).

Some users may wish to test a large set of annotations – for example, derived from pathways or cell types – conditional on a shared baseline model. We developed a fast score test for conditional heritability enrichment, graphREML-ST, that only requires fitting the baseline model once. This test runs in a few seconds, and does not even require access to the original summary statistics or LDGMs (**Online Methods**).

### Performance of graphREML in simulations

To evaluate the performance of graphREML and to compare it with that of S-LDSC, we simulated marginal association statistics using the LDGM precision matrices calculated from the European samples in the 1000 Genome project (**Online Methods**). Directly simulating summary statistics provides us with the flexibility to vary the sample size of the underlying association study. We used the S-LDSC baseline LD heritability model, and included 13 real functional annotations from the baselineLD model of the imputed SNPs on chromosome 1 (*p* = 513, 012) and 4 simulated annotations comprising randomly selected SNPs (**Online Methods, Supplementary Table** 1). We applied graphREML and S-LDSC to the simulated summary statistics and evaluated the bias and the variance of each method. In particular, we report their statistical relative efficiency (“RE”), defined as the ratio between the sampling variances of S-LDSC and graphREML. A RE value greater than one indicates graphREML is more statistically efficient than S-LDSC and vice versa for values less than one.

We found that both graphREML and S-LDSC produced unbiased enrichment estimates, but graphREML was much more precise with a RE of 2.47 averaged across annotations (**Figure** 1a, **Supplementary Figure** 1). For both methods, sampling variance is inversely correlated to sample size (**Supplementary Figure** 2); however, graphREML is more precise at any sample size, and its improvement upon S-LDSC is roughly equivalent to a two-fold increase in sample size (**Supplementary Table** 2). We analyzed random annotations of different size and connectedness, comprising 1% or 10% of either SNPs or LD blocks. For both methods, sampling variance was dependent on both factors (**Supplementary Figure** 3). The relative performance between graphREML and S-LDSC was similar when measured by mean square error, which accounts for both bias and variance (**Supplementary Table** 2).

**Figure 1.**
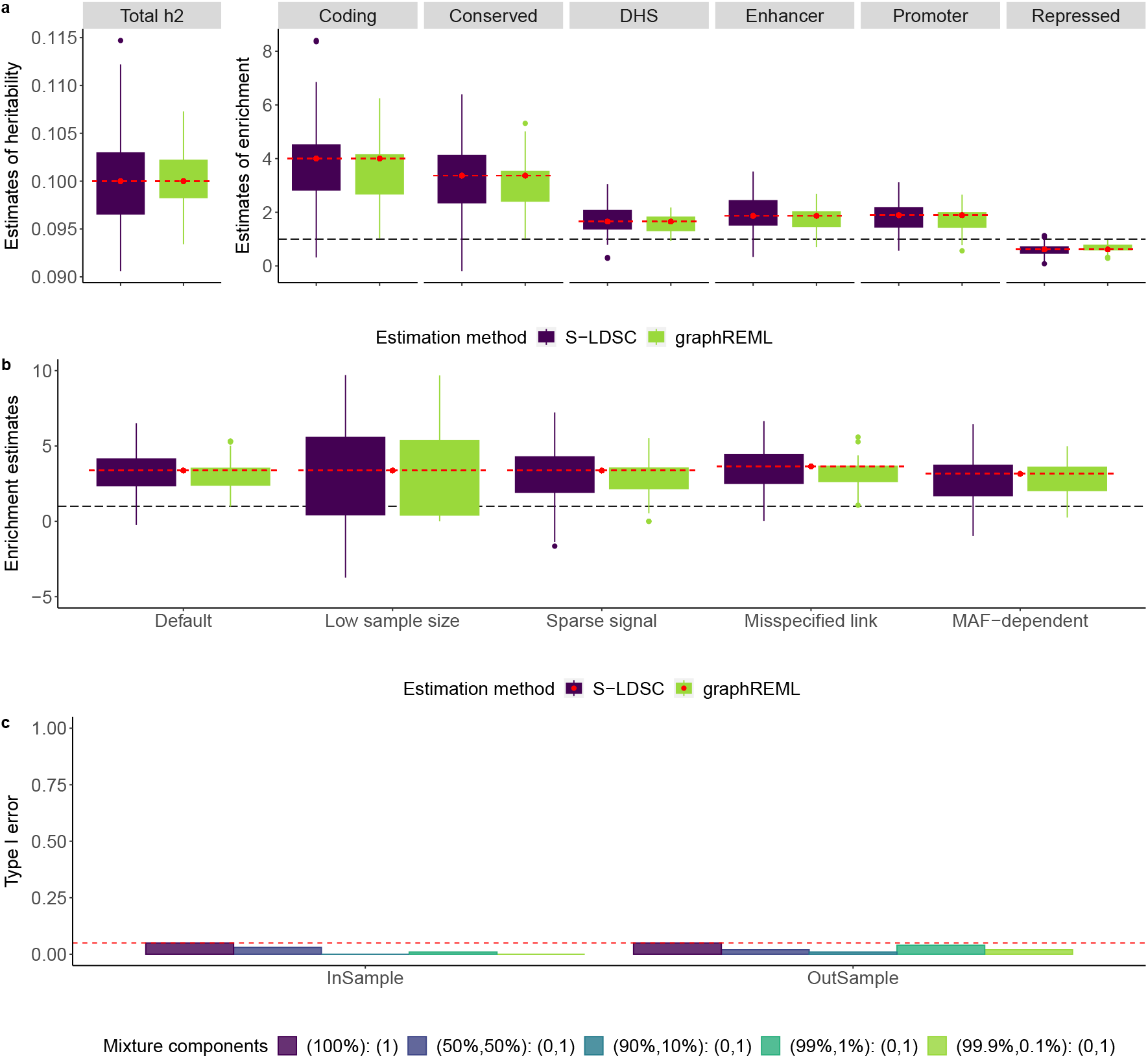
Performance of graphREML in simulation studies. Summary statistics are directly simulated based on the LDGM precision matrices on chromosome 1 from the Europeans in 1000 Genome (*p* = 513, 012). **a**. Comparison of heritability and enrichment estimates between S-LDSC and graphREML under the infinitesimal model, *n* = 100, 000. **b**. The enrichment estimates of conserved SNPs across different scenarios of model misspecifications and sample size. Low sample size: *n* = 10, 000; Sparse signal: 0.1% of SNPs are causal; misspecified link: use the max function to simulate genetic variances; MAF-dependent: assume the per-SNP heritability is proportional to (*f* _*j*_(1 − *f* _*j*_))^1+*α*^, where *α* = −0.25 and *f* _*j*_ is the allele frequency of SNP *j*. The results for the complete set of model misspecifications and for other functional annotations (*e*.*g*., coding) are reported in **Supplementary Table** 3-5. In panels **a** and **b**, the red dashed lines represent the true values of heritability enrichment; the black long dashed lines represent null or an enrichment of one. The box plots for the “Low sample size” setting in panel **b** are truncated due to the large variation of the estimates. **c**. Type I error rate of the joint enrichment estimates from graphREML. Y-axis is the proportion of true nulls that have been falsely rejected. The null annotation is DHS (18.9% of SNPs). InSample and OutSample indicate whether the LDGMs are matched with the summary statistics. The red dashed line is the level used for testing 0.05.

Misspecification of the random-effect model is a potential source of bias in heritability estimation. To test the robustness of graphREML, we varied the genetic architecture of a simulated phenotype in three ways (**Online Methods**). First, we simulated effect sizes under a sparse, non-infinitesimal distribution, varying the proportion of causal SNPs. Second, we generated summary statistics using several different link functions, with its inverse mapping from the annotations of a SNP to its heritability. Third, we explicitly modeled the MAF-dependent genetic architectures and varied the strength of the dependency^9^.

Under a sparse genetic architecture, both graphREML and S-LDSC remained unbiased and had higher sampling variance than that under an infinitesimal architecture. Across all settings of different mixture components and parameters, graphREML has a higher statistical efficiency, with an average RE of 2.73 compared to S-LDSC (**Figure** 1b, **Supplementary Figure** 4, **Supplementary Table** 3). Similarly, both methods were robust to the choice of link function and remain unbiased, but graphREML is more statistically efficient than S-LDSC, with an average RE of 2.54 across link functions (**Supplementary Figure** 5, **Supplementary Table** 5). In simulations involving MAF-dependent architecture, again, both methods were robust when we included MAF bins as binary annotations (as implemented in the baselineLD model^7^) (**Figure** 1b). This approach, with binary MAF-bin annotations, yielded more robust heritability estimates than the approach of using a single continuous-valued MAF annotation, likely because the former is nonparametric and imposes less constraint on the form of the relationship between allele frequency and effect size (**Supplementary Figure** 6, **Online Methods**). We did not detect any correlation between sparsity or the degree of MAF-dependency and the relative efficiency comparing graphREML with S-LDSC (**Supplementary Figure** 7, **Supplementary Table** 3-4).

We evaluated the calibration of our estimated standard errors. Under the null, we found that the jackknife-based significance for the conditional enrichment has well-controlled type I error rates (**Figure** 1c). We observed slightly inflated type I error rates for small null annotations, but only under the sparsest simulated architecture (**Supplementary Figure** 19), consistent with previous studies^33^. In non-null simulations, graphREML was well-powered (**Supplementary Figure** 18). We compared our jackknife approach with the Huber-White sandwich estimator (**Online Methods**). The inference results for the conditional enrichment coefficients are similar between using the jackknife estimator and the sandwich estimator (**Supplementary Figure** 20), but the jackknife estimator leads to more well-calibrated SE for the marginal enrichments than the sandwich estimator under sparse architectures (**Supplemetnary Figure** 8). Therefore, graphREML produces both estimators of SE but uses jackknife for testing by default.

A limitation of S-LDSC is that for individual SNPs as opposed to annotations, its per-SNP heritability estimates are unreliable, and in particular often negative. Weissbrod *et al*. proposed a procedure that led to well-calibrated per-SNP heritability estimates (which were used as valid prior causal probabilities in PolyFun), but the procedure requires re-fitting the S-LDSC after binning the SNPs, which is *ad hoc* and can be computationally intensive^20^. graphREML produces nonnegative per-SNP heritability estimates, which may be more reliable. We evaluated their calibration in our simulations involving different sample sizes and forms of model misspecification. We fit the graphREML model, used it to estimate the heritability of each SNP, and ranked SNPs by their estimated heritability. Then, we calculated the cumulative heritability explained by the top *x*%, for *x* ranging from 0 to 100, of variants in our list, and compared this curve with our estimates. These curves were highly concordant overall, though the degree of concordance is reduced when the genetic architecture is sparse, when sample size is low or when the genetic architecture is MAF-dependent (**Figure** 2, **Supplementary Figure** 10-11).

**Figure 2.**
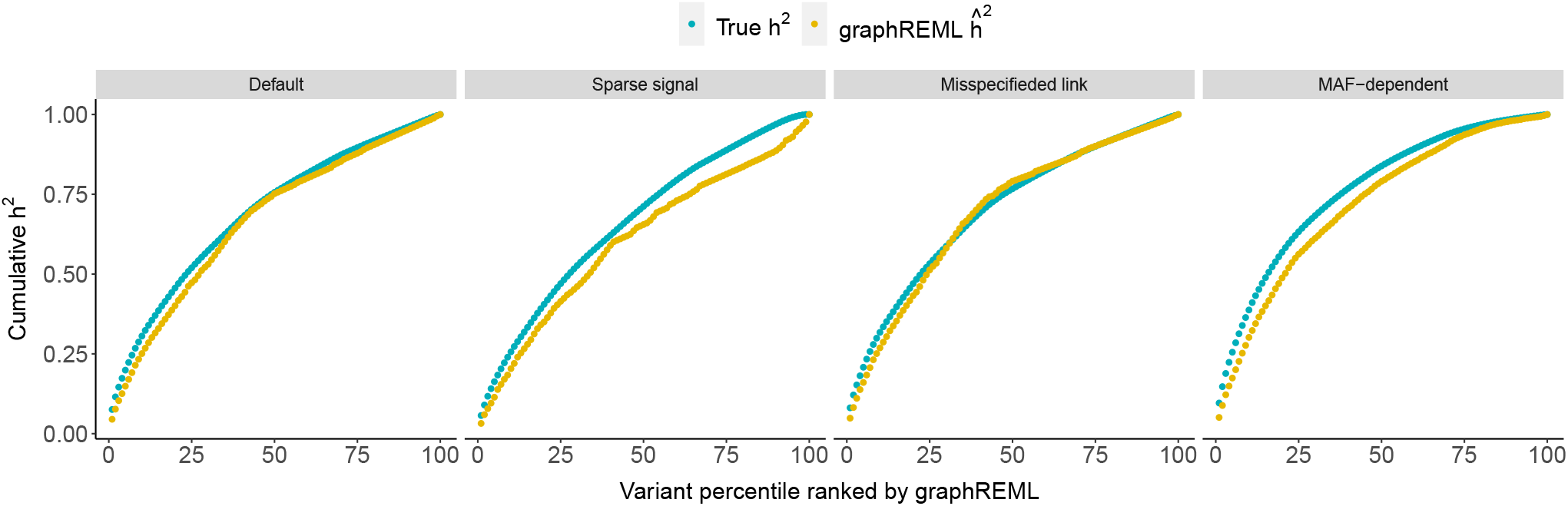
Calibration of per-SNP heritability from graphREML in simulations. Summary statistics are directly simulated based on the LDGM precision matrices on chromosome 1 from the Europeans in 1000 Genome (*p* = 513, 012). Each dot represent the estimated or true heritability from the top *k* percentiles of SNPs. Default is the infinitesimal model with *n* = 100, 000. Column panels represent different generative models or genetic architectures, similar to those defined in **Figure** 1b.

To further evaluate the calibration of per-SNP heritability, we regressed the estimated values onto the true values, constraining the intercept to be 0. The slope estimate from these regressions are close to 1, indicating a high degree of agreement between the estimated and true per-SNP heritability (**Supplementary Table** 6). We considered other approaches to assess the calibration of per-SNP heritability (**Online Methods**) and observed similar results (**Supplementary Figure** 9-11). These analyses indicates that for well-powered traits, variants with an estimated per-SNP heritability of some value *x* do indeed explain that much heritability on average.

Real summary statistics often contain a limited set of SNPs, for example the 1.1M HapMap 3 SNPs^34^. Missingness is potentially problematic in heritability enrichment analyses because when a missing causal variant in one annotation is in LD with a non-missing tag variant in a different annotation, its heritability might be misassigned. A particular advantage of S-LDSC is that it addresses this problem by explicitly modeling the LD of the “regression SNPs” via LD scores, which are computed based on a maximally comprehensive set of “reference SNPs”. Other methods for partitioned heritability estimation, such as RSS^30^ and GREML-LDMS^35^, cannot account for missingness or the mis-alignemnet between the set of variants with GWAS effect sizes and the set of variants with LD information. Our BaselineLD annotation matrices and LDGMs both contain a relatively comprehensive set of common SNPs; in particular, LDGMs contain most common SNPs in 1000 Genomes (MAF > 0.01; *p* = 8, 392, 958 for Europeans).

graphREML handles missingness in the summary statistics by assigning, for every missing SNP, a “surrogate marker” SNP in high LD (**Online Methods**). The heritability of the missing SNP is assigned to its surrogate marker appropriately. To test this approach, we simulated different degrees of missingness, and applied graphREML with and without surrogate markers. With surrogate markers, graphREML enrichment estimates were highly robust even when up to 90% of SNPs were missing, at which point it was strongly biased without surrogate markers (**Supplementary Figure** 12-13). Total heritability estimates were robust with up to 30% missingness, and they were downwardly biased (even with surrogate markers) when missingness was 40% or greater (**Supplementary Figure** 14).

### Methods comparison on UK Biobank phenotypes

On the basis of our simulation results, we expected that graphREML enrichment estimates would be concordant with those from S-LDSC on average, but that they would be less noisy, especially for traits with lower power. We analyzed UK Biobank summary statistics (average *n* = 451, 069 European-ancestry individuals) for 7 well-powered quantitative traits^36, 37^ as well as 11 less-well-powered disease phenotypes derived using the liability threshold family history model (“LTFH”)^4^ (**Supplementary Table** 7). We used a new set of LDGMs derived from UK Biobank data, closely matching the summary statistics; these LDGMs were highly accurate (**Supplementary Figure** 15). We applied both S-LDSC and graphREML to estimate the heritability enrichment of six selected annotations (coding, conserved, DHS, enhancer, promoter and repressed), in a joint analysis including the 96 annotations of the baselineLD model derived from the UK Biobank^8^.

Because both graphREML and S-LDSC were approximately unbiased in simulations, we expected that they produce concordant enrichment estimates on average across traits. We meta-analyzed 7 well-powered quantitative traits and the 11 disease traits, and found that indeed, the enrichment estimates were largely concordant between the two methods, and the estimates from graphREML are much less variable than S-LDSC (**Figure** 3a). Moreover, the enrichments of individual (*i*.*e*., as opposed to meta-analyzed) well-powered quantitative traits are similar as well (**Figure** 3b), For example, for height, coding variants had a heritability enrichment of 13.52 (*s*.*e*. = 2.47) with S-LDSC and 13.88 (*s*.*e*. = 1.83) for graphREML. The enrichment of variants in DHS are 3.56 (*s*.*e*. = 0.563) based on S-LDSC and 3.59 (*s*.*e*. = 0.214) based on graphREML.

**Figure 3.**
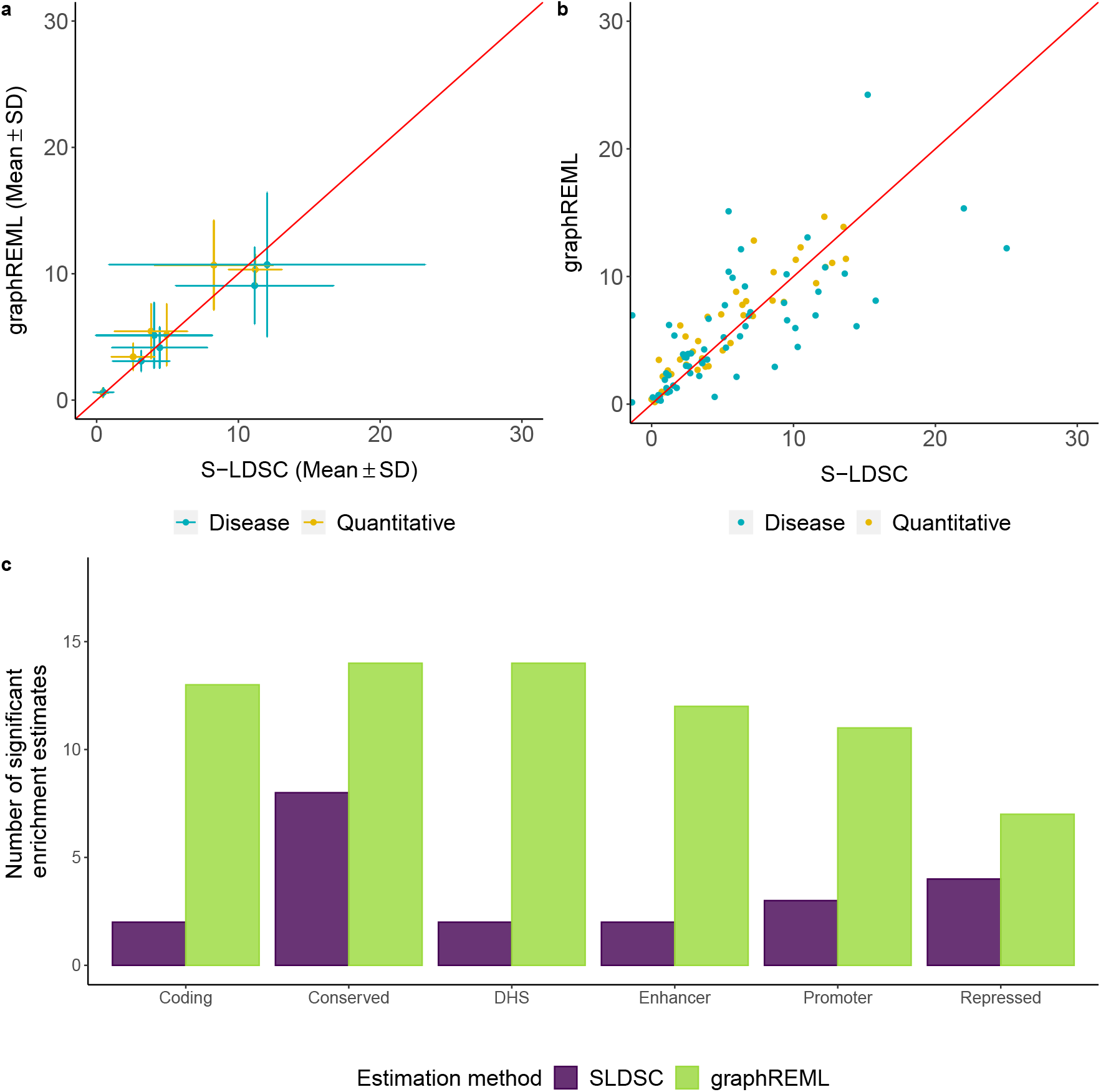
Comparison of marginal enrichment estimates from real trait analyses using graphREML vs. S-LDSC. Phenotypes are categorized into two groups: quantitative and disease, colored in yellow and blue respectively. The quantitative traits are generally better powered than the disease traits. Since the association statistics for the disease traits we use are based on the liability threshold model conditional on family history^4^, the disease traits are sufficiently well-powered as well. **a**. Marginal enrichment estimates from a meta-analysis of 18 traits based on the GWAS summary statistics in the UK Biobank. Error bars represent standard deviations (not standard errors) across traits. **b**. Marginal enrichment estimates for individual traits. The red reference line in panels **a** and **b** is the 45 degree line. **c**. Counts of significant enrichments identified by S-LDSC and graphREML across the 18 traits analyzed. Shown here are 81 significant trait-annotation pairs prioritized by graphREML vs. 32 pairs by S-LDSC. All significant enrichments identified by graphREML and S-LDSC have the correct direction (enrichment vs. depletion). The full set of enrichment estimates from the comparison are reported in **Supplementary Table** 8.

For the less-well-powered LTFH phenotypes, S-LDSC and graphREML still produced similarly concordant estimates on average, but for individual diseases, their estimates diverged (**Figure** 3b). For example, for cardiovascular disease, repressed variants had a heritability enrichment of 0.58 from S-LDSC and 0.59 from graphREML, but the standard error from S-LDSC is more than three times larger (*s*.*e*. = 0.247) than that from graphREML (*s*.*e*. = 0.082). Thus, this annotation would be identified as significantly depleted by graphREML but not by S-LDSC. Another example is prostate cancer, for which the enrichment of enhancer variants was estimated to be 6.9 from both S-LDSC and graphREML, but the standard error estimates were 4.11 from S-LDSC vs. 2.66 from graphREML.

More generally, graphREML better prioritizes the functional categories that are expected to be significantly enriched (or depleted) due to its better statistical efficiency (**Figure** 3c). For instance, graphREML identifies both DHS variants (×2.17, *p* = 4.42 × 10^−12^) and promoters (×3.47, *p* = 5.35 × 10^−8^) as highly significantly enriched for neuroticism. In contrast, S-LDSC produces noisy estimates for these two categories of SNPs – ×0.78 (*p* = 0.0724) for DHS and 0.51 (*p* = 0.57) for promoters, respectively. Across all trait-annotation pairs considered, 81 were statistically significant using graphREML vs. 32 using S-LDSC (**Supplementary Table** 8). Reassuringly, all of the significant discoveries identified from S-LDSC and graphREML have the expected directions of enrichment/depletion.

Next, we performed a secondary analysis to assess the impact of missing SNPs on graphREML. Many GWAS report summary statistics for HapMap3 SNPs only, or some other limited set of SNPs, potentially leading to bias. To evaluate the effectiveness of surrogate markers when missingness is more severe, we applied graphREML to the subset of HapMap3 SNPs for the traits we studied above. Despite having almost 89% of missingness in the summary statistics, the enrichment analyses using HapMap3 SNPs only produced estimates that were highly concordant with those from using the full set of SNPs in UK Biobank. (**Figure** 4a). Furthermore, we found that accounting for the missing variants led to improved power compared to ignoring them, although the improvement was modest due to the low level of missingness in the UKB summary statistics (**Figure** 4b).

**Figure 4.**
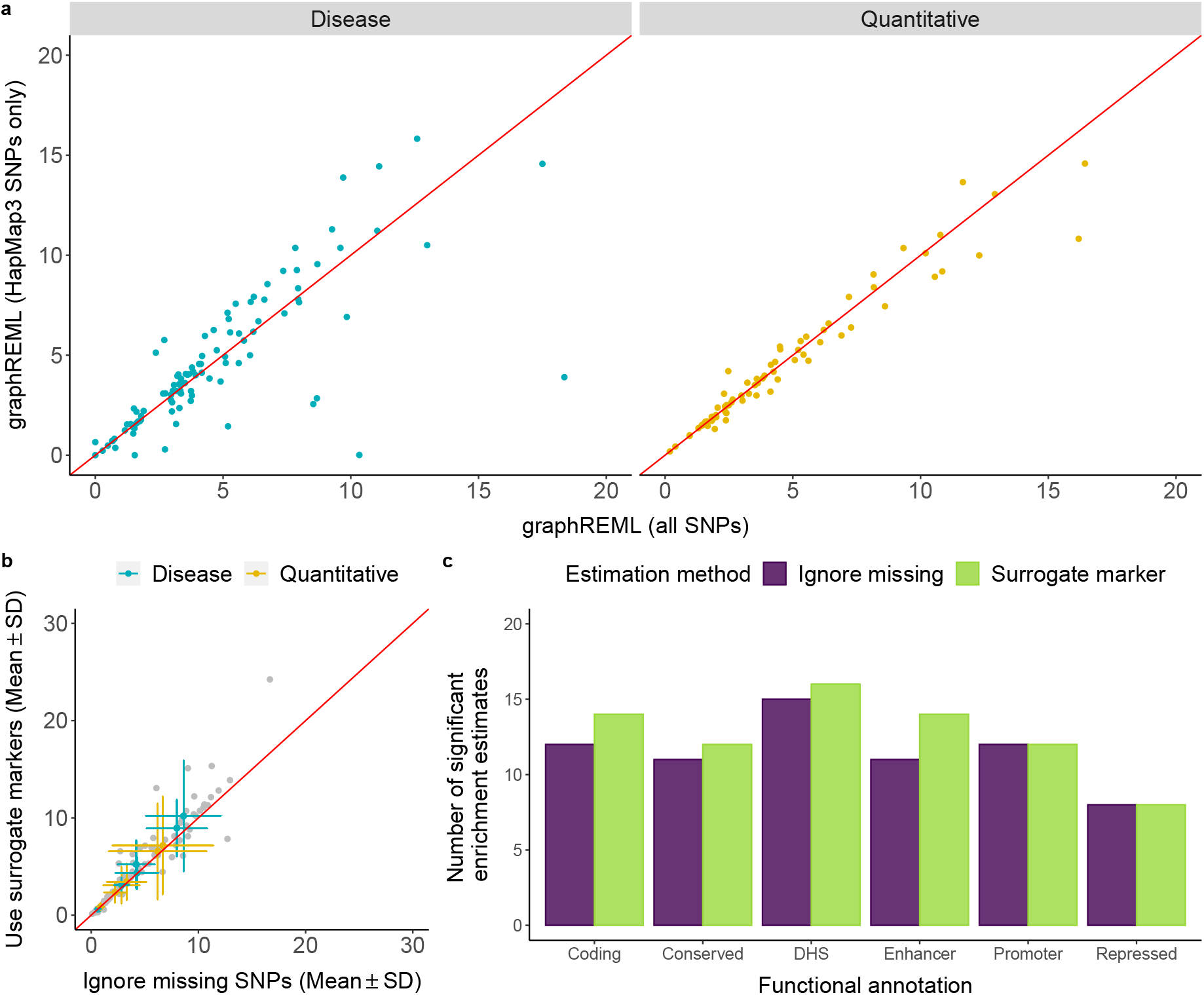
Robustness of graphREML in the presence of missing SNPs. The set of phenotypes is the same as those used for method comparison in **Figure** 3. **a**. Comparison of the enrichment estimates when the full set of SNPs (x-axis) vs. HapMap3 only (y-axis) are included in the summary statistics. Approximately 89% of the SNPs in the full summary statistics are not HapMap3 SNPs. **b**. Enrichment estimates from meta-analyses of 18 traits from GWAS in the UK Biobank when ignoring the missing SNPs (x-axis) vs. when accounting for the missing SNPs using surrogate markers (y-axis). Approximately 17.82% of the SNPs in the UKB summary statistics are missing in the LDGMs (UKB-based). Gray dots represent the enrichment estimates for specific trait-annotation pairs. Phenotypes are categorized into two groups: well-powered traits and low-powered traits, colored in yellow and blue respectively. The lines represent SD within each group. The red reference line is the 45 degree line. **c**. Counts of significant enrichment/depletion identified by graphREML when accounting for the missing SNPs via surrogate marker vs. ignoring the missing SNPs.

Lastly, we analyzed the UK Biobank traits using non-UK Biobank LDGMs derived from 1000 Genomes European individuals. These enrichment estimates were concordant with those involving UKB-derived LDGM precision matrices, although power was reduced (**Supplementary Figure** 17). These results support the use of graphREML with out-of-sample LDGMs, and highlight the broad utility of graphREML for publicly available GWAS summary statistics.

### Validation of graphREML in non-UK Biobank datasets

Most GWAS involve genotype data that is not publicly available, and they release summary association statistics but no in-sample LD information. Our method is derived under a model where the genotype matrix is random, with a population LD matrix that could potentially be estimated *out-of-sample*. We evaluated the performance of graphREML in such datasets, comparing its results with those obtained within UK Biobank. We identified non-UK Biobank, European-ancestry summary statistics for 12 of the traits analyzed above (average *n* = 235, 331; **Supplementary Table** 9). We additionally analyzed Biobank Japan summary statistics for 19 of the traits analyzed using European individuals and the LDGM precision matrices derived from East Asians in the 1000 Genome (average *n* = 91, 045; **Supplementary Table** 10). Most of these summary statistics were limited to HapMap3 SNPs (around 11% of those contained in the LDGM).

Enrichment estimates were concordant between UK Biobank and non-UK Biobank summary statistics for the same traits (**Figure 5a**-b, **Supplementary Figure** 21, **Supplementary Table** 11), both across the 6 annotations analyzed above and across a larger set of annotations having > 5% of SNPs (**Online Methods**). Well-powered quantitative traits had strongly concordant estimates; for example, the enrichment estimates for height based on UKB and non-UKB Europeans have a correlation of *r*^2^ = 0.973 with a mean enrichment across annotations of 4.89 and 4.99 based on UKB and non-UKB European GWAS, respectively. Less-well-powered disease traits had less concordant estimates, consistent with sampling error, but they were still concordant after meta-analyzing across traits (**Figure** 5b). These results also support the application of graphREML to estimate heritability enrichment in a sample which is potentially different from the LDGM sample.

**Figure 5.**
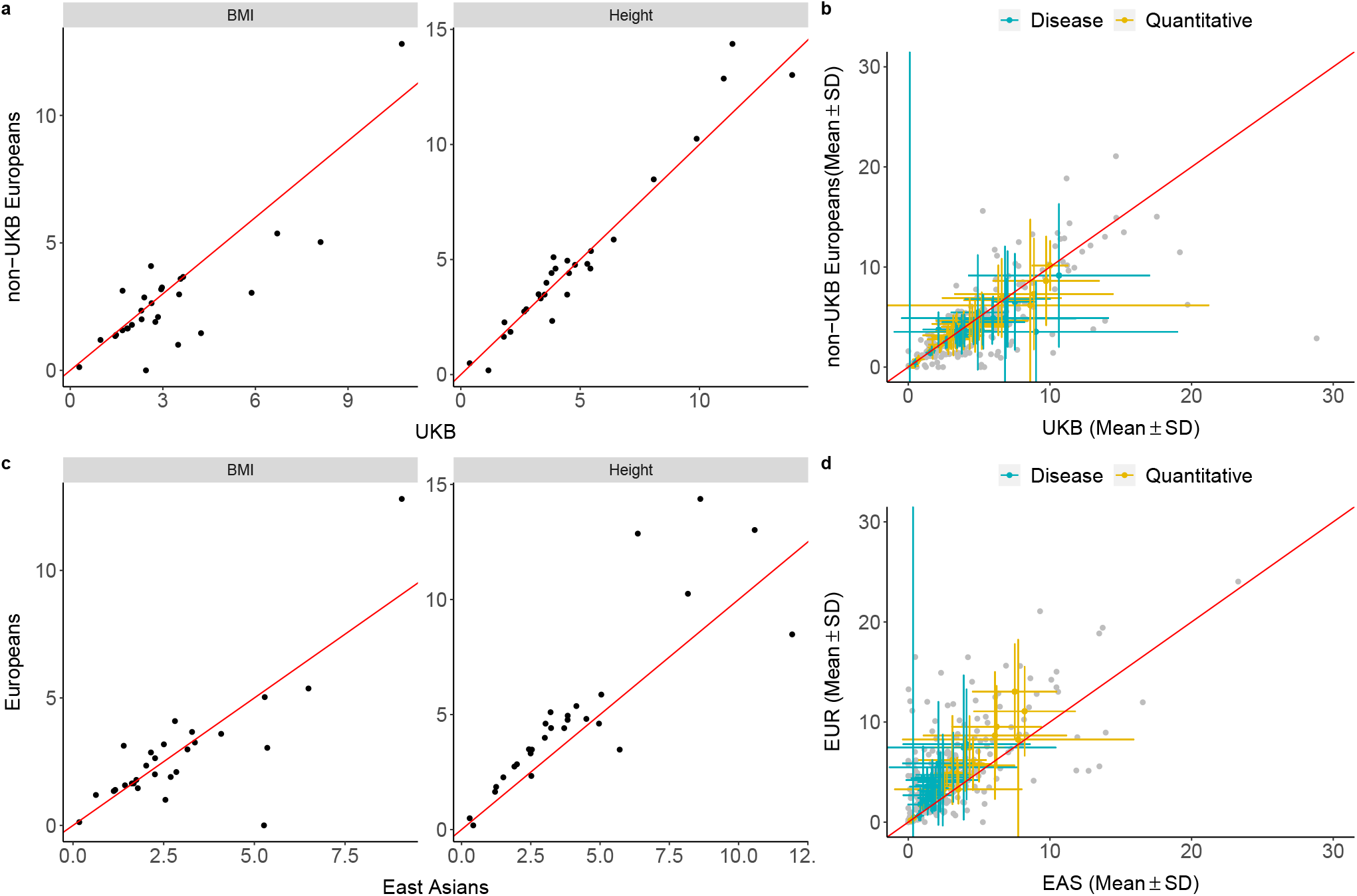
Validation of graphREML in non-UK Biobank datasets. **a**. Enrichment estimates from applying graphREML to GWAS for BMI and height in UKB or non-UKB Europeans. LDGM precision matrices derived from the UKB are used. **b**. Meta-analysis results from applying graphREML to 12 traits with both UKB and non-UKB summary statistics. The red reference line is the 45 degree line. The full set of enrichment estimates are reported in **Supplementary Table** 11. **c**. Enrichment estimates from applying graphREML to BMI and Height GWAS based on Biobank Japan (BBJ) and UKB. LDGM precision matrices derived from UKB and 1000 Genomes East Asians are used with UKB and BBJ GWAS respectively. **d**. Meta-analysis results from applying graphREML to 11 traits with both UKB and BBJ summary statistics. The red reference line is the 45 degree line. The full set of enrichment estimates are reported in **Supplementary Table** 13.

Finally, we analyzed summary statistics from Biobank Japan in conjunction with LDGMs derived from East Asian individuals in 1000 Genomes. For most traits, estimates were concordant with those based on UK Biobank, including height and BMI (**Figure** 5c-d). For all seven blood traits or hematopoietic phenotypes in our study, enrichments in Biobank Japan were consistently smaller than those derived from European ancestry GWAS (**Supplementary Figure** 22). We observed similar results when comparing between East Asians and non-UKB Europeans (**Supplementary Figure** 23, **Supplementary Table** 12-13).

To remove the potentially large effect of the MHC/HLA region on the enrichments of hematopoietic phenotypes, we reran the analyses with the variants in the MHC/HLA region excluded. The enrichment estimates were largely concordant when we included vs. excluded the HLA region. (**Supplementary Figure** 24); the cross-ancestry comparison had a similar pattern with the HLA region excluded (**Supplementary Figure** 25). Together, these estimates are consistent with previous studies finding a high cross-population genetic correlation between European and East Asian populations^38–40^. They support the notion that different ancestry groups have differences in their allele frequencies and LD patterns, leading to different GWAS results, but that the underlying biology (in particular, function architecture) is mostly shared.

### A fast test for heritability enrichment

In many studies, a large number of annotations are tested for heritability enrichment, conditional on the same baseline annotations^5, 41–43^. Using a Wald test to obtain the significance of a conditional enrichment requires refitting graphREML multiple times, with the annotation of interest swapped in and out. This is analogous to the heritability enrichment analyses of specifically expressed genes (SEG) using S-LDSC, where a separate regression is ran for each SEG annotation and inference is performed on the regression coefficient on the SEG annotation^5^. While the regression step of S-LDSC is fast, estimating the enrichment of a new annotation requires calculating a new set of LD scores first, which is not computationally trivial.

We derived a fast test for heritability enrichment, graphREML-ST, that circumvents the need of refitting graphREML or computing the LD scores for each new annotation, conditional upon a shared null model (**Online Methods**). The main advantage of this procedure is that it is based on a score test, and hence only requires running graphREML once to fit the null model (**Online Methods**). The test is computationally efficient, with runtime linear in the number of markers (**Supplementary Notes**).

We evaluated the performance of the score test in simulations (**Online Methods**). We first assessed the type I error rate and the power of the score test. We found that the false positive rate is well-controlled across different genetic architectures with varying degree of polygenicity (**Supplementary Table** 14). Moreover, the score test has sufficient statistical power to detect true enrichment under a range of realistic generative models (**Supplementary Table** 15). We also compared the inference results based on the Wald test versus the score test from the real-trait enrichment analyses of 8 quantitative and 12 disease phenotypes in the UK Biobank (**Online Methods**). Reassuringly, we observed a high degree of concordance between the two set of inference results, with a Kendall’s coefficient of concordance greater than 0.87 and 0.82 for the marginal and joint enrichment, respectively (**Supplementary Table** 16). Taken together with the simulation results validating the Wald test (shown above), the agreement between the two tests lends support to using the score test as an optimal and robust approach to identify relevant annotations that are significantly enriched for a disease or trait.

This test is highly convenient for users because it allows them to test their new annotation for enrichment against traits for which we have already run graphREML. They can do so without re-fitting graphREML to the summary statistics; they do not need to download LDGMs or even the original summary statistics. We have released the null fit from applying graphREML to the baseline LD annotations for a set of complex traits and diseases in the UK biobank (see data availability).

### Application of graphREML to the Abstract Mediation Model

The abstract mediation model (AMM) is a model for the distance-dependent relationship between trait-associated variants and the genes that might mediate their effects. Under the assumption that all variant effects are mediated by some nearby gene, it quantifies the fraction of heritability that is mediated by the closest, second-closest, or kth-closest genes. It leverages the proximity of SNPs to genes belonging to an enriched gene set to partition gene-mediated heritability.

AMM was previously paired with S-LDSC for estimation, because it requires an enrichment model that can handle overlapping annotations. As a result, its estimates are noisy. Consequently, the estimates have the limitations that they have low statistical efficiency, and are derived based on a linear assumption about the effect of an annotation on per-SNP heritability. To address these limitations, we apply graphREML to AMM to estimate the fraction of heritability mediated by the *k*-th nearest genes (**Online Methods**). Notably, we adopt a flexible mapping to relax the linear assumption on the relationship between the annotation values of a SNP and its per-SNP heritability. We also allow the background heritability of a SNP (*i*.*e*., per-SNP heritability if no nearby genes lies in the gene set) to depend on its functional annotations. Denote by *p*^(*k*)^ the proportion of the total heritability mediated by the *k*-th nearest genes. To increase power and to ensure the stability of our estimates of the *p*^(*k*)^ estimates, we bin the gene proximity annotations, and perform meta-analyses across traits (**Online Methods**).

In simulations, we verified that the *p*^(*k*)^ estimates are approximately unbiased under different sample sizes and misspecified genetic architectures (**Supplementary Table** 17). We observed slight downward bias for the true non-null bins and upward bias for the true null bins. We emphasize that such biases are expected as we use a non-negative estimator of *p*^(*k*)^ with the implicit constraints that *p*^(*k*)^ *>* 0 and ∑_*k*_ *p*^(*k*)^ = 1. We next applied graphREML in conjunction with AMM to estimate *p*^(*k*)^ for the same set of traits as we used to validate graphREML using real-trait data. We found that the *p*^(*k*)^ estimates are largely consistent with those reported in the previous study, with the closest and the 2nd closest gene mediating approximately 22.9% and 9.9% of the SNP-heritability in meta-analyses (**Figure** 6a, **Supplementary Table** 19). Notably, our estimates are more precise than those reported in the original AMM, even with fewer traits used for meta-analyses^29^. For instance, the standard errors of *p*^(1)^ and *p*^(2)^ from meta-analyzing 15 traits using the graphREML enrichments are 2.91% and 2.32%, whereas those from meta-analyzing 47 traits using the S-LDSC enrichments are 6.37% and 3.79%, respectively. We observed large variation in the *p*^(*k*)^ estimates across traits; in particular, these estimates are more precise for well-powered and polygenic traits (**Figure** 6b-c, **Supplementary Table** 18). For well powered traits, graphREML can produce precise estimates of *p*^(*k*)^ for each *individual* trait, whereas such estimates were not reported in Weiner *et al*. due to lack of power.

**Figure 6.**
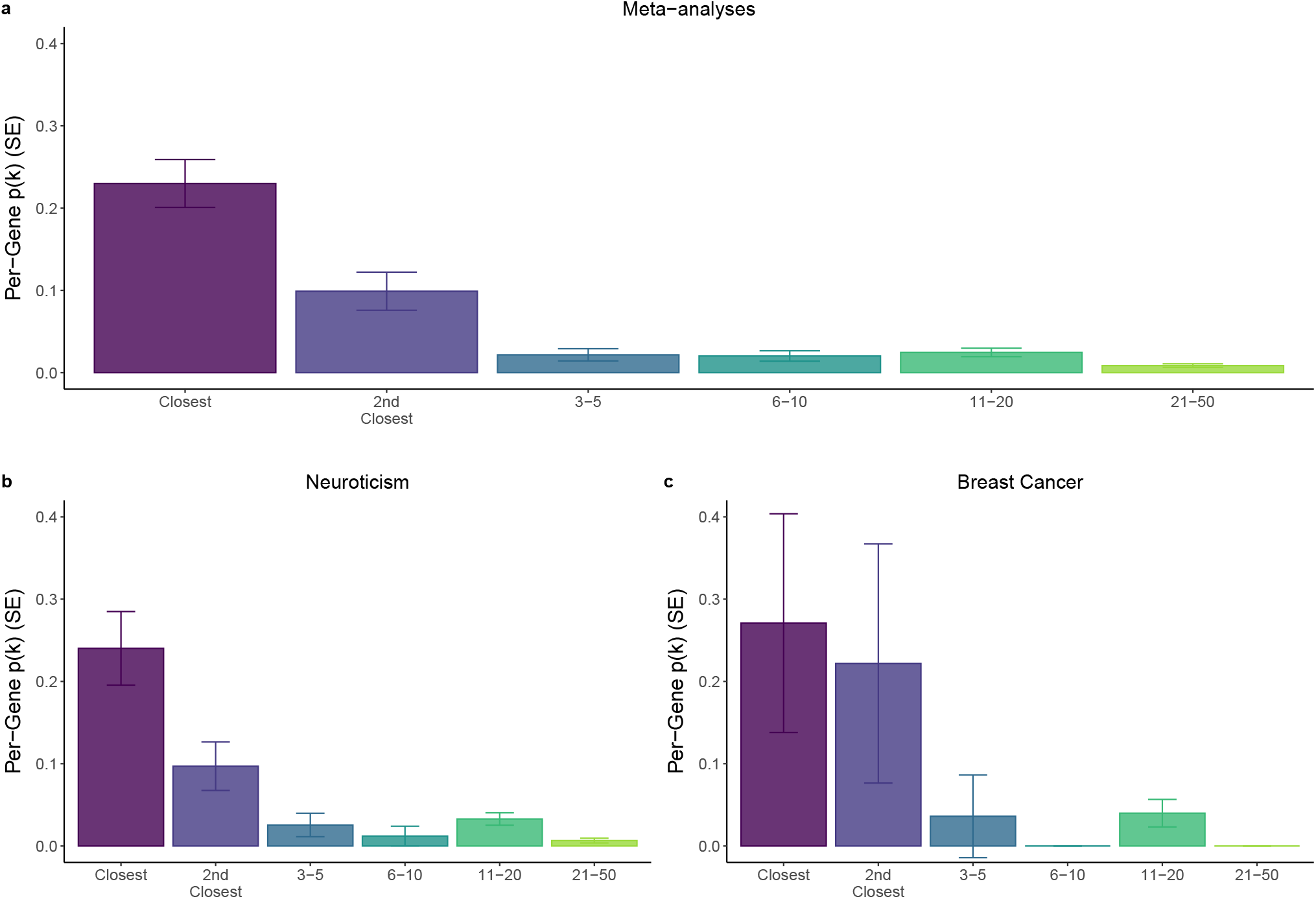
Application of graphREML to the Abstract Mediation Model (AMM). Fraction of mediated heritability across gene-proximity bins estimated with the constrained gene set (pLI≥0.9). The estimate of *p*^(*k*)^ is the average for genes in that bin; per-bin *p*^(*k*)^ multiplied by the number of genes in the bin, summed across bins, equals 100% of heritability. Error bars represent standard errors from jackknife. **a**. Meta-analyzed estimates across traits, weighted by the precision of excess heritability (*τ*_*A*_). **b** and **c**. *p*^(*k*)^ estimates for two individual traits: neuroticism and breast cancer. The numerical results for all panels are reported in **Supplementary Table** 18-19.

## Discussion

Heritability enrichment analysis has been one of the most valuable approaches to understand genetic architecture and to link functional genomic datasets with disease genetics. Here we proposed a new summary statistics-based approach and demonstrated through extensive simulations and real-trait analyses that compared to existing methods, graphREML has significantly improved statistical efficiency and power for enrichment analyses, and is robust to mismatches between the summary statistics and LD.

Model-based estimates of heritability and heritability enrichment can be biased due to misspecification of the assumed heritability model^7, 15, 28, 32^. graphREML can be used to fit essentially any heritability model, notably including the baselineLD model, which includes a set of annotations that are designed to account for LD-dependent and frequency-dependent architecture, and which has been extensively validated using S-LDSC. These phenomena should affect graphREML and S-LDSC similarly, and indeed, both methods produce concordant estimates under the baselineLD model (Figure 3a). A completely different approach is to eschew the use of any heritability model, treating genetic effects as fixed; this approach is impervious to misspecification-related bias, but it has not been successfully applied to heritability partitioning with overlapping annotations^44, 45^.

Two other types of model misspecification are non-infinitesimal effect sizes and misspecified link functions. Bayesian methods such as RSS-NET explicitly models the null effects through its specification of the prior^46^. In contrast, graphREML assumes a Gaussian likelihood, similar to that of GCTA. Though our simulation results support the application of graphREML to non-infinitesimal architectures, further research is needed to study the impact of a sparse architecture on enrichment estimation. We proposed using a non-negative link function to map the annotation vector of a genetic marker to its per-SNP heritability. While our results suggest that the softmax function leads to well-calibrated estimates and is generally robust to model misspecification, future research is needed to improve the modeling of per-SNP heritability, in particular the form of the link function. For example, one can develop a data-adaptive procedure to select the most appropriate link systematically.

Another important source of bias when estimating heritability is assortative mating^47^, which causes long-range correlations between trait associated variants, magnifying their marginal effects. Assortative mating is expected to affect total heritability estimates more strongly than it does enrichment estimates. However, cross-trait assortative mating^48^ would affect graphREML-estimated enrichments to the extent that the pattern of enrichment varies between the traits under assortment. This bias is expected to be similar for any heritability estimator that does not model assortative mating explicitly.

The likelihood of the marginal summary statistics we use in graphREML has been used in other methods as well. One such method is High-Definition Likelihood (“HDL”)^17^, which estimates the genetic correlation between two traits with higher statistical efficiency than cross-trait LDSC. A possible extension of graphREML would be to partitioned genetic correlation. Another is “Regression with Summary Statistics (RSS)^30^,” which estimates heritability but does not allow for overlapping annotations; moreover, it operates on a limited set of SNPs due to computational limitations. We recently developed a likelihood-based estimator, HEELS, which is approximately equivalent to individual-level REML estimator, again operating on a limited set of SNPs and not allowing for annotation overlap. A key difference between the two methods is that HEELS requires *in-sample* LD whereas graphREML can incorporate LDGM precision matrices that are estimated either in-sample or out-of-sample^25^. For precise total heritability estimation, we recommend using HEELS when in-sample LD information is available.

It is worth noting that because graphREML cannot distinguish polygenic effects from confounding due to population stratification, it requires the S-LDSC intercept as an input to correct for confounding. Nevertheless, we observed largely consistent heritability enrichment estimates when we ignored population stratification (*i*.*e*., fixing the intercept at 1 instead of the S-LDSC estimated intercept) (**Supplementary Figure** 26). Another limitation of graphREML in comparison with S-LDSC is that it is much slower, with a runtime on the order of hours vs. minutes (**Supplementary Table** 20). This makes it less well-suited for exploratory analyses involving hundreds of traits and annotations. graphREML-ST can alleviate this limitation to the extent that when a large number of annotations need to be tested conditional on a shared set of annotations, one only needs to run graphREML once for the null fit and apply score test to the new annotations, which only takes a few seconds. Another potential approach to improve the graphREML runtime would be stochastic optimization, where each update is computed from a subset of the genome.

Increasingly, genomic datasets resolve subtle differences between cell types, between nearby SNPs, across time points, and within tissues. With such an increasing resolution, these datasets will require powerful methods to prioritize disease-relevant mechanisms. The statistical efficiency of graphREML can be leveraged, in conjunction with high-resolution functional data, to identify highly specific features of disease biology.

## Online Methods

### Statistical model

Let **y** be a length-*n* vector that denotes the phenotypes of *n* samples. Denote by **X** ∈ ℝ^*n*×*p*^ the genotype matrix of *n* individuals based on *p* markers or SNPs. We standardize **X** and **y** such that the variance of the phenotype is 1 and the variance of each marker-specific genotype vector is 1. We adopt the standard assumptions of genetic association modeling, and use an additive genetic model for the phenotypes as **y** = **X***β* + *ε*, where both **X** and *β* are assumed to be random. We define the population LD matrix as **Σ** ≡ 𝔼(**X**^⊤^**X***/n*), and we assume the true effect sizes are drawn from *β* ~ *N*(0, **D**(*θ*)), similar to Yang *et al*.^14^, *θ* is the set of parameters that determine the genetic architecture of a trait. For example, *θ* can include the total heritability of a trait; it can also include the set of enrichment coefficients for the functional annotations, *i*.*e*., *τ*. The diagonal elements of **D**(*θ*) are the per-SNP heritability, which we model using the link function, *g*(·). We use the softmax by default in graphREML: for SNP *j*, we allow for a non-linear relationship between the annotation values of a SNP and its per-SNP heritability value, as described above,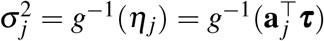. We assume that the individual-specific noise is *i*.*i*.*d*., following 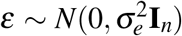. Under this random-design random-effect model, the marginal association statistics, **z**, is normally distributed with mean zero, and variance approximately equal to *n***ΣD**(*θ*)**Σ** + **Σ** (**Supplementary Notes**,^49^).

Maximizing the likelihood of this model requires computationally expensive operations involving the LD matrix Σ. We propose to approximate the likelihood using the LDGM precision matrix^26^, *P*, which is a sparse matrix whose inverse approximates Σ. We define transformed Z-statistics, *i*.*e*., 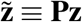, whose likelihood is:

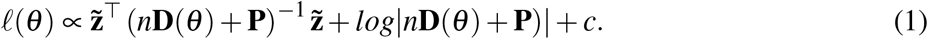

The graphREML estimator is defined as 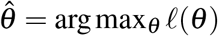.

### Estimation

We use the Newton-Raphson algorithm to maximize the likelihood function (1) and we exploit the sparse representation of **Σ**^−1^ with the LDGM precision matrices. We iteratively update our estimate of the parameters as the following,

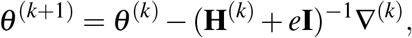

where ∇^(*k*)^ and **H**^(*k*)^ are the gradient and Hessian of the likelihood function evaluated at the current estimate of the parameters *θ* ^(*k*)^. *e* is some small-valued number that is added to the diagonal of the Hessian matrix to prevent singularity in estimation.

Let **M**(*θ* ^(*k*)^) = *n***D**(*θ* ^(*k*)^) + **P**. At each iteration, we first perform a Cholesky factorization of the matrix **M**(*θ* ^(*k*)^), which is feasible and computationally tractable due to the sparsity of **P**. Specifically, we use the sparse matrix operations in MATLAB to efficiently obtain the Cholesky factors. The diagonal elements of **D**(*θ*) correspond to the true SNP-specific genetic variance, which are normalized such that they summed up to the true total heritability. Denote by 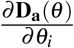 the diagonal matrix where the diagonal elements are the partial derivatives of the per-SNP heritability with respect to the parameters, 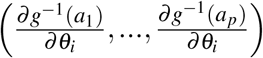. The gradient is:

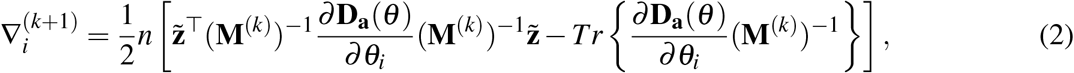

where we have used **M**^(*k*)^ to denote **M**(*θ* ^(*k*)^) for simplicity of notation, and *i* indexes the parameters. The second term is computationally intensive to evaluate; for this, we calculate the sparse inverse subset^50^ using the *suitesparse* library in MATLAB^51, 52^.

For the Hessian matrix, we apply the trace trick to compute the expected value of a quadratic form and approximate the expected information using the observed information, similar to Loh et al.^16^ (**Supplementary Notes**). This leads to an approximation of the Hessian as,

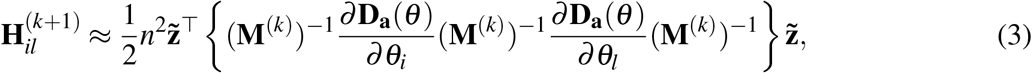

where *i* and *l* index the parameters. We compute the inverse **M**^−1^ in both equation (2) and (3) using the Cholesky factors described above. All computations are performed at LD-block level (the LDGM precision matrices are provided by LD block as well), which can be parallelized. We use the trust-region algorithm to control the step size of each update in a principled way, and employ an adaptive bound on the maximum change at each iteration (see Algorithm 1 in the **Supplementary Notes**). This allows us to balance between convergence speed and robustness of the updates. We apply other techniques to improve the computational efficiency of our algorithm. Notably, out calculations critically rely on the sparsity of the LDGMs, as we use sparse matrix operations for matrix multiplication, division, log-determinant and inverse (**Supplementary Notes**). It is possible to have multiple SNPs on the same LDGM node. graphREML chooses just of the SNPs with available summary statistics for a given node, and sum up the estimated per-SNP heritabilities across the SNPs that are assigned to the same node.

### Standard error calibration

We estimate the standard error of our estimates using an approximate jackknife estimator. Specifically, we compute a set of leave-one-LD-block-out scores (the score is the gradient of the log-likelihood function) at the optimum, after the Newton Raphson algorithm has completed. This amounts to performing one more NR update using all but one LD block. Such an approximation is appropriate because our variance estimator is most exact when it is evaluated at the true parameter value, and we expect our estimates to be close to the optimum upon completion of the Newton updates. We then use the empirical variance of these leave-one-LD-block-out parameter estimates as the jackknife covariance of the conditional enrichment coefficients, *τ*. graphREML also produces the Huber-White sandwich estimator, where we plug in the empirical covariance of the scores across the LD blocks as the weight, sandwiched by inverse of the naive variance estimator which is the inverse of the negative Hessian. Both the jackknife estimator and the sandwich estimator of SE lead to well-controlled type I error rates except under very sparse architectures, *e*.*g*., 0.1% causal (**Supplementary Figure** 19-20).

For inference on the marginal enrichment, we adopt an approach similar to S-LDSC, testing the significance of the difference rather than the ratio between the partitioned heritability in vs. out of an annotation. We apply the Delta method to obtain the asymptotic variance of the enrichment test statistics *D*_*k*_, which are functions of the conditional enrichment estimates *τ*. By default, graphREML reports inference results based on the jackknife estimator of SE, as it is slightly more conservative for the marginal enrichment under sparse architectures (**Supplementary Figure** 8). We apply the standard Benjamini-Hochberg procedure to correct for multiple hypothesis testing when prioritizing trait-annotation pairs, such that the false discovery rate (FDR) is less than 5%.

### Accounting for missing SNPs

In practice, the sets of SNPs that are present in the summary statistics are almost always different from the set of SNPs with annotation and/or the LD information available. To address such missingness issue, graphREML assigns to each missing SNP a surrogate marker, selected as the non-missing SNP which has the highest LD with the missing SNP, and uses these surrogate markers in heritability enrichment estimation. Note that this procedure accounts for the set of SNPs that we have annotation and LD information for but are absent in the summary statistics. We cannot model or include SNPs with association statistics available but no annotation or LD information in the graphREML estimation. This latter type of missingness is not concerning because in practice, the set of SNPs that are included in the LDGM precision matrices and with annotation information is usually a superset of the common variants with available association statistics. We note two important points about merging between variant-level data in real data analyses due to the imperfect alignment of the SNPs across datasets. First, we explicitly model the covariance of the effects of SNPs that overlap between the summary statistics and the LDGM nodes in modeling the per-SNP heritability. Due to the surrogate markers we assign to the missing SNPs, it is possible that one LDGM node is linked with multiple SNPs. In this case, we aggregate the per-SNP heritability of all of the SNPs on a given node. Second, it is possible for multiple SNPs to be assigned to the same LDGM node (*i*.*e*., if their LD is almost perfect), in which case we retain all of the variants with available annotation information as opposed to randomly selecting one.

To evaluate the impact of SNP missingness in real data analyses, we first compared the enrichment estimates from applying graphREML with versus without accounting for missing SNPs, using summary statistics in the UK Biobank. Since the proportion of missing SNPs is small in UK Biobank (especially if we use the LDGM precision matrices derived from the UK Biobank), we next applied graphREML to the subset of summary statistics that overlap with the HapMap3 SNPs. This led to approximately 1.2 million variants in the summary statistics and close to 89% missingness.

### Simulation studies

We simulated summary statistics using the *simulateSumstats* function in the LDGM package (see Code Availability). To simulate the association statistics directly, we drew marginal effect size from the multivariate normal, *N* (0, *n***ΣD**(*θ*)**Σ**), where **Σ** is the inverse of the LDGM precision matrix inferred from European individuals in the 1000 Genome or in the UK Biobank. We used the imputed common SNPs on Chromosome 1 (*p* = 513, 012 for 1000 Genome or *p* = 504, 907 for UK Biobank). We varied the form of **D**(*θ*) to (mis)specify different generative models and architectures. The diagonal elements of **D**(*θ*) correspond to the true SNP-specific genetic variance, which are normalized such that they summed up to the true total heritability. Under the infinitesimal model, **D**(*θ*) = *diag*(*h*^2^*/p*,…, *h*^2^*/p*); for sparse architectures, we simulated the joint effect sizes from a mixture of normal components, one of which is null (*i*.*e*., point mass at zero); for frequency-dependent architectures, we assumed that the genetic variance of SNP *j* was proportional to a function of its allele frequency, 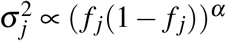, where *α* determines the strength of the dependency^9, 28^. We considered two alternative links for SNP-specific heritability, the exponential function,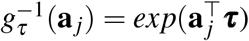, and the ReLU activation function, 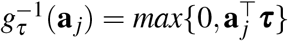 (Note that the max function is not invertible, but we keep using the *g*^−1^ notation to be consistent with the GLM literature, where link refers to the mapping from the expected response to a linear combination of predictors).

We computed the LD scores using the LDGM precision matrices by taking the sum of the squared correlations between two SNPs. This ensures that the set of variants used for graphREML and S-LDSC is closely aligned. To further increase comparability between graphREML and S-LDSC in simulations, we included SNPs with large *χ*^2^ in S-LDSC at the regression step (since graphREML accounts for all SNPs regardless of their effect sizes) even though by default, S-LDSC removes SNPs with *χ*^2^ greater than 80.

We also fixed the intercept of graphREML at the intercept estimated by S-LDSC such that confounding related to population stratification is adjusted in the same way between two methods. The 14 functional real functional annotations we included in the simulation studies are coding, conserved, enhancer, DHS, DHS peaks, promoter and repressed, along with their flanking regions (<500kb). We chose these annotations because they are well-known and studied; they are also well-powered (*i*.*e*., the annotations are not too small). In addition, we simulated 4 random annotations, which comprise 1% of the SNPs, 10% of the SNPs 1% of the LD blocks and 10% of the LD blocks. To account for frequency-dependent architecture in estimation, we either incorporated a single continuous-valued MAF annotation or a set of 10 binary MAF bins (same as the baselineLD model).

### Real trait analyses

We analyzed a diverse set of GWAS summary statistics downloaded from different sources (see URLs). For method comparisons, we applied graphREML and S-LDSC to estimate the heritability enrichment of 7 well-powered quantitative traits – height, BMI, red blood count (RBC), monocyte count (Mono), platelet count (Plt), years of education, and neuroticism, using the publicly available summary statistics^36, 37^. We also applied the two methods to the summary statistics of 11 less-well-powered disease phenotypes – Alzheimer’s disease (AD), bowel cancer, breast cancer, cardiovascular diseases (CAD), chronic obstructive pulmonery diseases (COPD), depression (DEP), hypertension (HTN), lung cancer, Parkinson’s disease (PD), prostate cancer and type II diabetes (T2D). The summary statistics for these traits were derived using the liability threshold family history model (LTFH)^4^.

For validation analyses, we applied graphREML to European-ancestry summary statistics for which UKB was not the major source of GWAS sample (average *n* = 235, 331; **Supplementary Table** 9). We also analyzed a set of summary statistics that are based on GWAS of East Asians (*n* = 91, 045; **Supplementary Table** 10). These summary statistics were identified based on their availability and to maximize overlap with traits we used for method comparison (**Supplementary Table** 7).

### Score test for inference on joint enrichment

We derived a score test to circumvent the need of refitting graphREML for every new annotation, as long as the set of baseline annotations to be conditioned on is the same. Below we outline the key steps of the score test procedure. Further details, such as its derivation, the intuitions and the computational aspects of the test are provided in the **Supplementary Notes**.

1. Run graphREML to fit the null model (*i*.*e*., only including the baseline annotations), and store two variant-wise statistics. (These values roughly correspond to the gradient and Hessian of the likelihoods, with respect to the per-SNP heritability. The exact definitions of these values are provided in the **Supplementary Notes**.)
2. Perform the score test
  a. Construct the score statistic using the SNP-specific values stored in Step 1, along with the new annotation to be tested. The score statistics for the test of a single annotation can be written as,

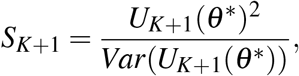

where *U*_*K*+1_(*θ* ^*^) is the score for the new annotation aggregated from all markers, and *Var*(*U*_*K*+1_(*θ* ^*^)) is the jackknife variance estimator (see details in **Supplementary Notes**).
  b. Compute the empirical variance of the score statistic using jackknife, leaving one LD block at a time. We account for the uncertainty in the parameter estimates of the null model (**Supplementary Notes**).
  c. Compute the p-value by comparing the normalized score statistic against the chi-squared distribution (with one degree of freedom if only one annotation is tested).
3. Compare the significance levels across the set of annotations tested and control for multiple testing.

This test is performed using an efficient block jackknife procedure, such that it accounts for the uncertainty in the parameter estimates from the null model (**Supplementary Notes**).

To evaluate the type I error rate and power of the score test, we simulated quantitative traits under different genetic architectures, using the real LDGM precision matrices derived from the UK Biobank, along with the same set of annotations as those used in the validation analyses (*i*.*e*., coding, conserved, DHS, enhancer, promoter and repressed). The joint effect sizes are drawn from a mixture of normal components, one of which is null (*i*.*e*., point mass at zero); we varied the proportion of null variants from 0% to 99.9% and normalized the total heritability to be 0.1 in all simulation settings. We assumed the true non-null annotations are coding and conserved. For type I error rate, we ran score test on promoter and repressed, conditional on all the other annotations. Note that our null model excludes the annotations to be tested for type I error, but includes the non-null annotations due to potential overlaps between the non-null and the null annotations (*e*.*g*., coding and promoter have large overlaps). For the power analyses, we varied the true enrichment of the non-null annotations, but largely kept them at realistic values. We referenced the published meta-analyses results from Finucane *et al*.^3^ for the estimated enrichment of coding and conserved SNPs from real traits. Their estimates from the meta-analyses are 7.124(0.842) and 13.318(1.503) for coding and conserved respectively. Alpha level is set to 0.05 for all of our tests.

To assess the concordance between the inference results from the Wald test and those from the Score test, we compared the enrichment estimates and their p-values for the same set of annotations and real traits as those used in the validation analyses. For clarity, we call the set of annotations we are interested in testing their enrichment the “new annotations”, and the set of annotations we want to condition on the “baseline annotations”. For the marginal enrichment, the p-values of the Wald test are based on fitting graphREML to the set of baseline annotations and new annotations jointly, after which we extract the p-values for the marginal enrichments of the new annotations; the p-values of the Score test are based on fitting graphREML to the all-one annotation first and then applying the score test to each of the new annotations separately in turn. For the joint enrichment, the p-values of the Wald test are based on fitting graphREML to the baseline annotations plus a new annotations, one at a time, and then extracting the p-values for the new annotations; the p-values of the Score test are based on fitting graphREML to the baseline annotations first and then applying the score test to each of the new annotations separately in turn.

### Applying graphREML to Abstract Mediation Model (AMM)

The abstract mediation model (AMM) characterizes the distance-dependent relationship between trait-associated variants and the genes that mediate their effects. In particular, it quantifies the fraction of heritability mediated by the closest, second-closest, or *k*th-closest genes, and leverages the proximity of SNPs to genes that belong to an enriched gene set to partition gene-mediated heritability.

Let 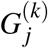 denote the *k*th closest gene to SNP *j* and let 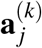 be an indicator *i*.*e*., binary annotation, of whether 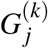 is in the gene set of interest, *A*. We model the heritability of SNP *j* mediated by its *k*th nearest gene as the following,

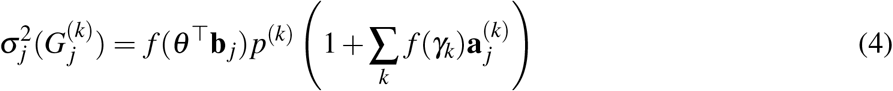

where *f* (·) is some smooth and non-negative function we choose for estimation, analogous to the inverse link above, *e*.*g*., softmax. For clarity, we use separate notations – *θ* and *γ* as parameters, **b** _*j*_ and **a** _*j*_ as annotation values – for the baseline and the *k*th nearest gene annotations, respectively.

We define the excess per-SNP heritability mediated by the nearest genes (in gene set A) as *τ*(*A*) ≡ ∑_*k*_ *f* (*γ*_*k*_). In other words, the excess heritability explained by a SNP with its closest *k*th gene in A, scaled by its background heritability, is *p*^(*k*)^*τ*(*A*). Summing up the mediated heritability across all nearest genes considered, we have the model for the expected per-SNP heritability *j*,

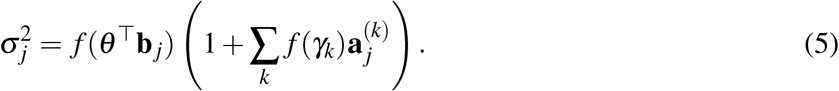

Similar to Weiner *et al*., we bin the gene proximity annotations to increase power. Let *q* denote a gene bin and let *n*_*q*_ denote the number of genes aggregated together for bin *q*. We estimate the fraction of per-SNP heritability mediated by the nearest genes in gene set A as 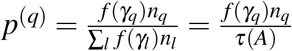. For meta-analyses, we weigh each trait by its total heritability before obtaining the average *p*^(*k*)^ across traits. We define the average mediated heritability enrichment of a gene set as 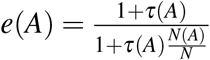, where *N*(*A*)*/N* is the fraction of genes in set A.

Our model differs from the original AMM model in two main ways. First, we allow the baseline annotations of a SNP (**b** _*j*_) to affect its “background heritability” or the per-SNP heritability if none of its nearby genes lies in the gene set. More specifically, S-LDSC models the heritability contributed from the baseline annotations and the excess heritability mediated by genes in the gene set additively,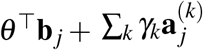; graphREML, on the other hand, assumes a multiplicative model and enables the “interaction” between the baseline annotations and the nearest gene annotations, 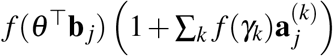. Second, we apply a non-negative link *f* (·) to ensure the validity of our per-SNP heritability estimates. In other words, S-LDSC accounts for the AMM annotations as 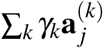 whereas graphREML uses 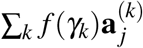. Our definition of enrichment is similiar to Equation 6 in Weiner *et al*. but differs slightly in that we assume *τ*(*A*) has been scaled by the background heritability at the SNP level, *i*.*e*., divided out by *f* (*θ* ^⊤^**b** _*j*_) as in Equation (4).

In simulations, we generated GWAS Z-scores using the *simulateSumstats* function in the LDGM package (see Code Availability). The true per-SNP heritability was defined using the following generative link function,

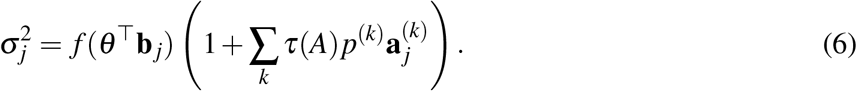

We started with the set of “baselineLD minus” annotations which exclude annotations that control for genic elements relevant to constrained genes, such as conservation, minor allele frequency, and ancient sequence annotation. Out of these 66 annotations, we randomly selected four baseline annotations to assign heritable signals (non-zero *θ* elements; these values are set to [1, 2, 1.5, 2.5]). We assume the fraction of mediated heritability across the four nearest genes as *p*^(*k*)^ = [0.4, 0.3, 0.2, 0.1]. The total excess per-SNP heritability of the enriched gene set A is *τ*(*A*) = 2. We applied graphREML to AMM and estimated the heritability mediated by the constrained genes. We varied the sample size to be 10^4^, 10^6^ and 10^8^, and generated the effect sizes under three levels of polygenicity, assuming 99.9%, 99%, 90% of the variants are null. Under each of these six settings, we repeated the simulations 30 time and ran graphREML using the full set of baseline annotations and the AMM link function in equation (4). Because we observed numerical stability issues due to the exponential terms, we modified our link function to address these overflow issues by implementing a piece-wise version of the softmax (**Supplementary Notes**).

For real-trait analyses, we applied AMM to the GWAS summary statistics for the same set of traits as we analyzed before, including both well-powered quantitative traits and disease traits with less power. We used the LDGM precision matrices derived from the UK biobank. We used highly constrained genes (pLI > 0.9) which are intolerant of heterozygous loss-of-function variation (see Weiner *et al*.) as our enriched gene set. To increase the power of our *p*(*k*) estimates, we binned annotations in the 3rd through 5th, 6th through 10th, 11th through 20th, and 21st through 50th nearest genes. We performed meta-analyses across traits by taking the ratio of the weighted averages,

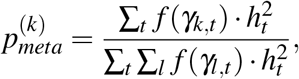

where the weights are the trait-specific total heritability. To obtain standard errors on the estimates, we use the jackknife values of *γ*_*k,t*_ to compute the jackknife estimates of 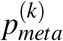. The standard error is computed as the standard deviation of these jackknife estimates, multiplied by square root of the number of LD blocks.

## Data availability

The baselineLD annotations can be downloaded on Google Cloud (https://storage.googleapis.com/broad-alkesgroup-public-requester-pays/LDSCORE/baselineLF_v2.2.UKB.tar.gz). The constrained gene sets can be downloaded from the AMM Github repository (https://github.com/danjweiner/AMM21/blob/main/AMM_genesets/AMM_gs_constrained.txt). LDGM precision matrices derived from the 1000 Genome are available from Zenodo (https://doi.org/10.5281/zenodo.8157131).

## Code availability

Our method (graphREML) has been implemented as an open-source package, written primarily in Matlab, available on Github at https://github.com/huilisabrina/graphREML. We also used the open-source LDGM package and S-LDSC package, available on Github at https://github.com/awohns/ldgm and https://github.com/bulik/ldsc.

**References**

## Acknowledgements

We are very grateful to Alkes Price and Samuel Kou for their helpful discussions and feedback. We thank Dan Weiner for his assistance to Tushar Kamath with the application of graphREML to AMM. We thank the participants of the individual in the UK Biobank. This work was supported by grants R35-CA197449, U19-CA203654, R01-HL163560, U01-HG012064, and U01-HG009088 (to X. L.) and by grant R35 GM155278 (to L.O.).

## Author contributions statement

H.L., T.M., L.O. and X.L. conceived and designed the experiments. H.L. performed the experiments and the statistical analyses. H.L. and L.O. wrote the manuscript with the participation of R.M. and X.L. L.O and X.L. supervised the project.

