## Supplementary Figures for "Improved heritability partitioning and enrichment analyses using summary statistics with graphREML"

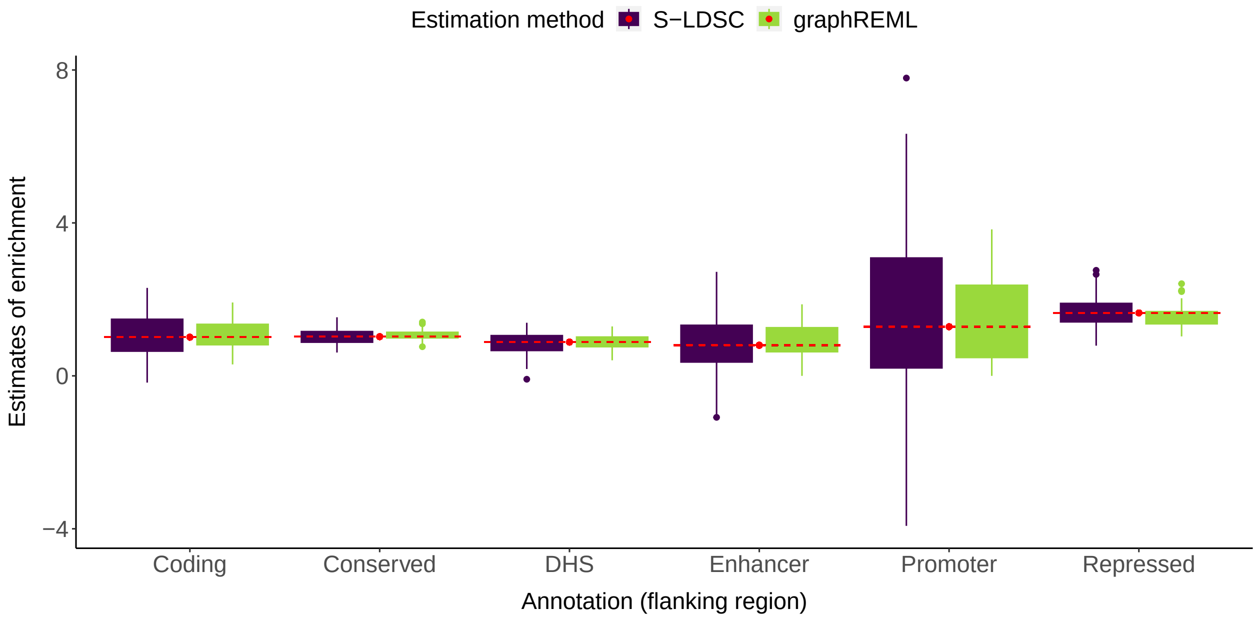


**Supplementary Figure 1. Comparison of enrichment estimates for the flanking region of the functional categories.** Summary statistics are directly simulated based on the LDGM precision matrices on chromosome 1 from the Europeans in 1000 Genome ($p = 513,012$, $n = 100,000$). The boxplots show the empirical distributions of the enrichment estimates from S-LDSC or graphREML. The red dashed lines represent the true values of heritability enrichment.

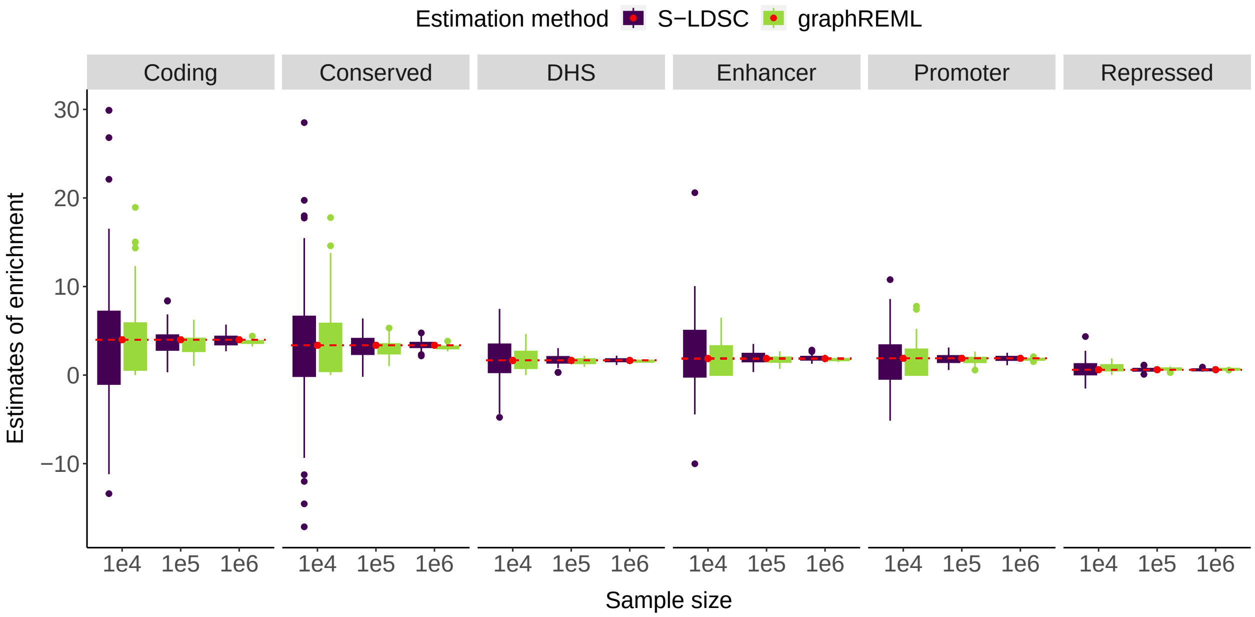


**Supplementary Figure 2. The impact of sample size on the enrichment estimates from S-LDSC vs. graphREML.** Summary statistics are directly simulated based on the LDGM precision matrices on chromosome 1 from the Europeans in 1000 Genome ($p = 513,012$). The boxplots show the empirical distributions of the enrichment estimates from S-LDSC or graphREML. The column panels correspond to different functional annotations. The red dashed lines represent the true values of heritability enrichment, which differ across annotations. The numerical values are reported in **Supplementary Table** 2.

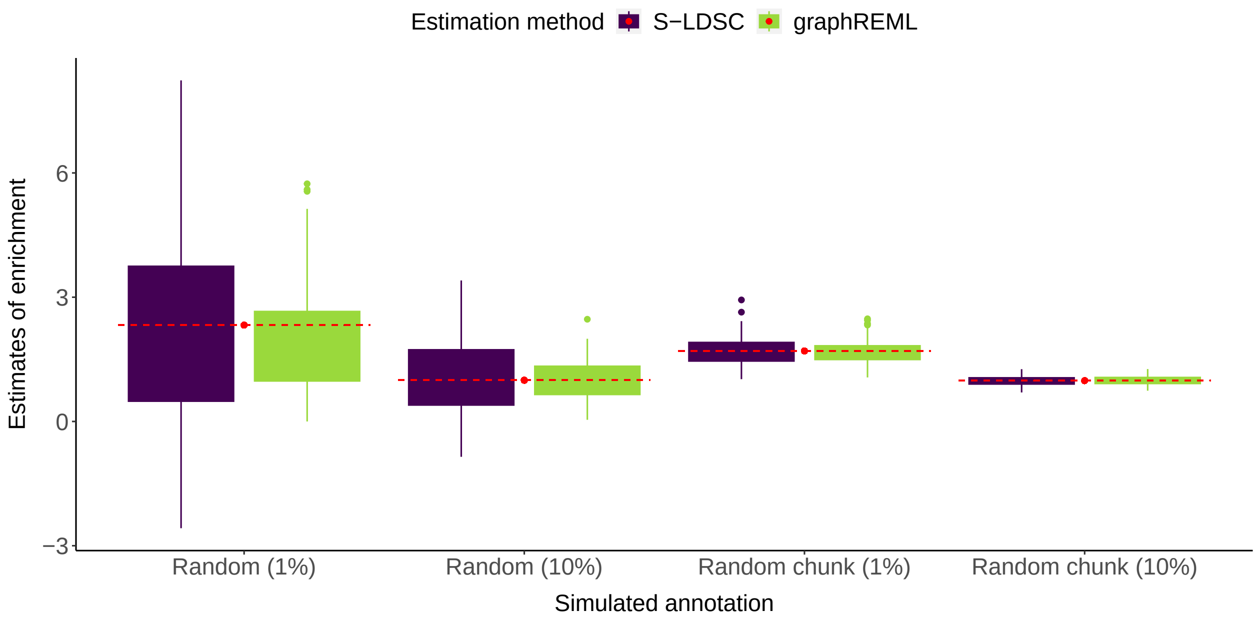


**Supplementary Figure 3. Comparison of enrichment estimates for the random annotations.** Summary statistics are directly simulated based on the LDGM precision matrices on chromosome 1 from the Europeans in 1000 Genome ($p = 513,012$, $n = 100,000$). The boxplots show the empirical distributions of the enrichment estimates from S-LDSC or graphREML. "Random (1%)" corresponds to the annotation that randomly assigns 1 to 1% of the SNPs on the genome; "Random chunk (1%)" corresponds to an annotation where a random continuous region with the size of 1% of the genome is assigned 1. The red dashed lines represent the true values of heritability enrichment.

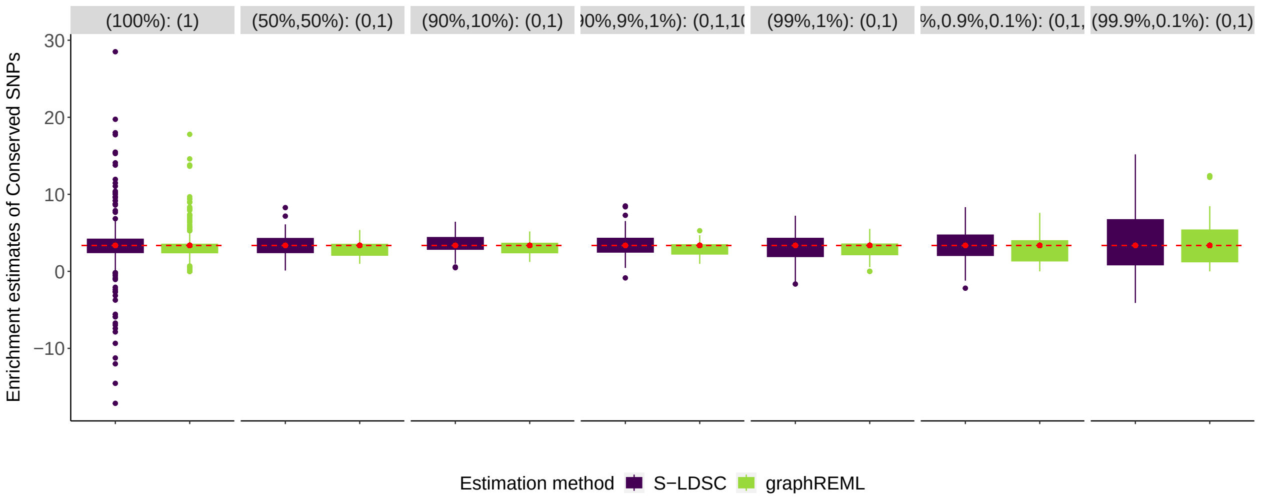


**Supplementary Figure 4. Robustness of enrichment estimates under model mis-specifications: varying causal signal sparsity**. Summary statistics are directly simulated based on the LDGM precision matrices on chromosome 1 from the Europeans in 1000 Genome ($p = 513,012, n = 100,000$). The column panels represent different genetic architectures or mixture of causal effects, denoted as (component weights): (variance of the normal components). For example, (99%, 1%): (0,1) signifies that 99% of the SNPs are null and 1% of markers have their effect sizes drawn from $N(0,1)$. The red dashed lines represent the true enrichment value.

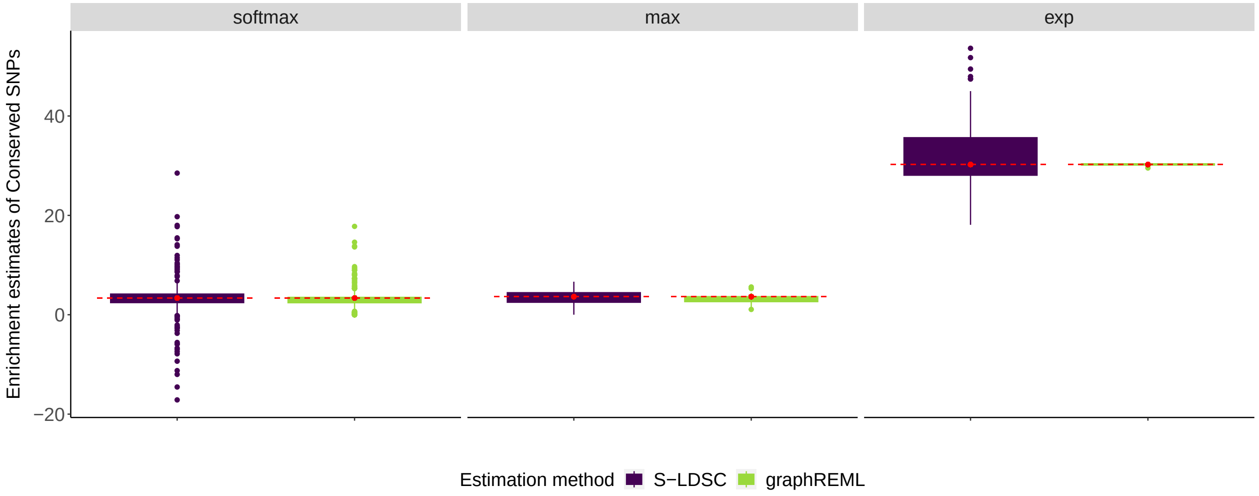


**Supplementary Figure 5. Robustness of enrichment estimates under model mis-specifications: varying generative link function**. Summary statistics are directly simulated based on the LDGM precision matrices on chromosome 1 from the Europeans in 1000 Genome ($p = 513,012, n = 100,000$). The column panels represent different link functions used to generate the causal effect size variances. The default option of softmax is used to produce all graphREML estimates. The red dashed lines represent the true enrichment value.

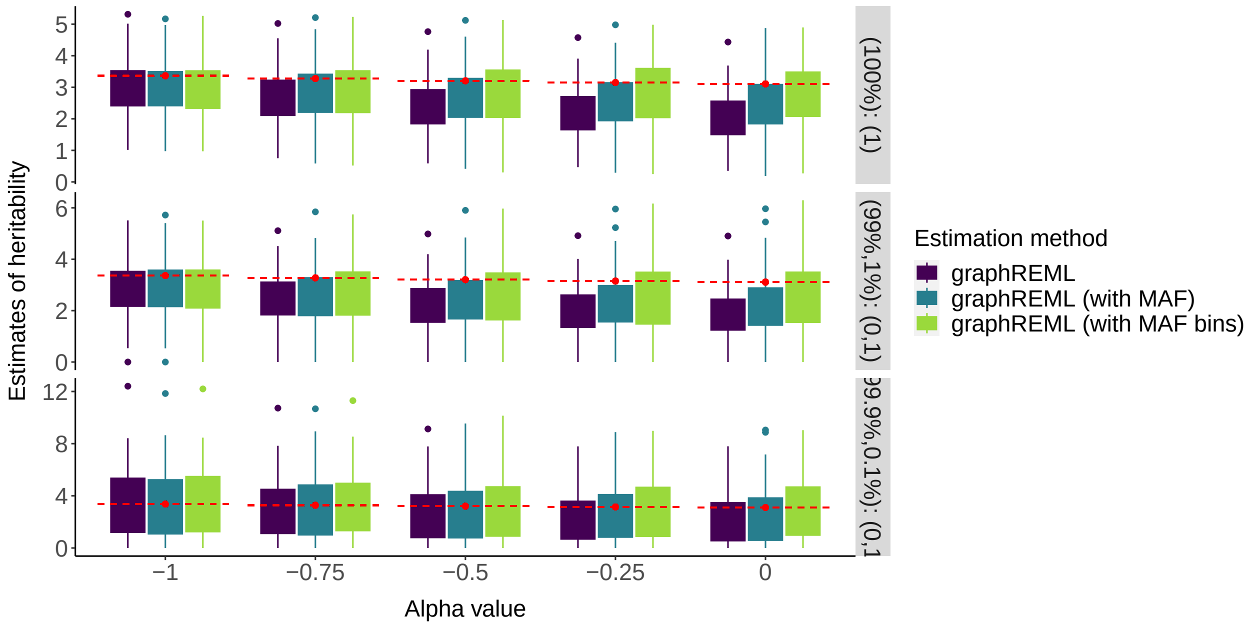


**Supplementary Figure 6. Robustness of enrichment estimates under model mis-specifications: varying frequency-dependent architecture**. Summary statistics are directly simulated based on the LDGM precision matrices on chromosome 1 from the Europeans in 1000 Genome ($p = 513,012, n = 100,000$). Row panels represent different genetic architectures or mixture of causal effects, denoted as (component weights): (variance of the normal components). For example, (99%, 1%): (0,1) signifies that 99% of the SNPs are null and 1% of markers have their effect sizes drawn from $N(0,1)$. X-axis correspond to different frequency-dependent architectures. The ALpha value determines the strength of the dependency[1]. For example, $\alpha = -1$ corresponds to the GCTA model and $\alpha = -0.25$ corresponds to the LDAK model. The red dashed lines represent the true heritability value of 0.1. graphREML: estimation without inclusion of any MAF-related annotations; graphREML (with MAF): estimation including MAF as a single continuous annotation; graphREML (with MAF bins): estimation that accounts MAF via 10 binary annotations for the 10 deciles.

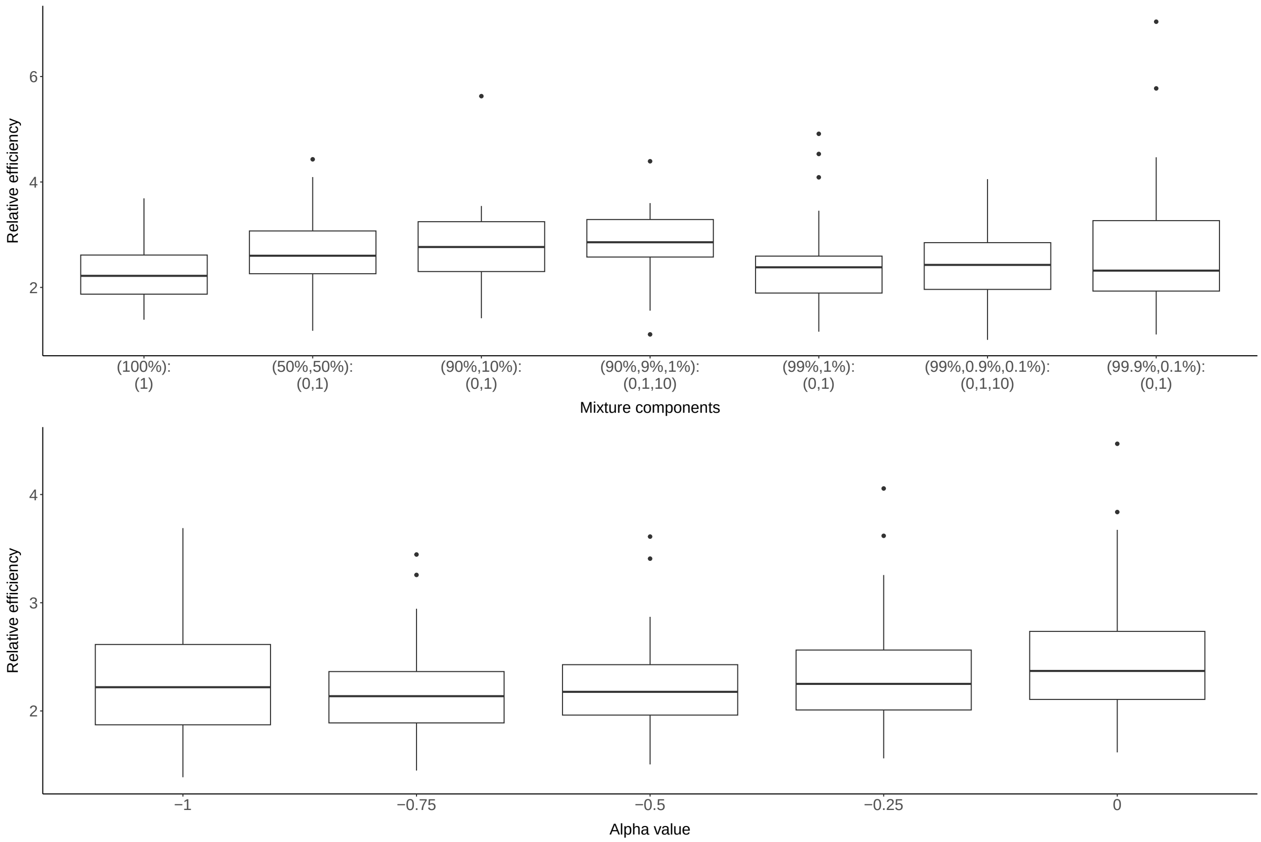


**Supplementary Figure 7. Comparison of relative efficiency across different genetic architectures.** The boxplots show the distributions of RE across parameters. The top panel corresponds to RE with varying mixture components; the bottom panel corresponds to RE with varying degrees of frequency-dependence. Numerical values of these results are provided in **Supplementary Table** 3 and 4.

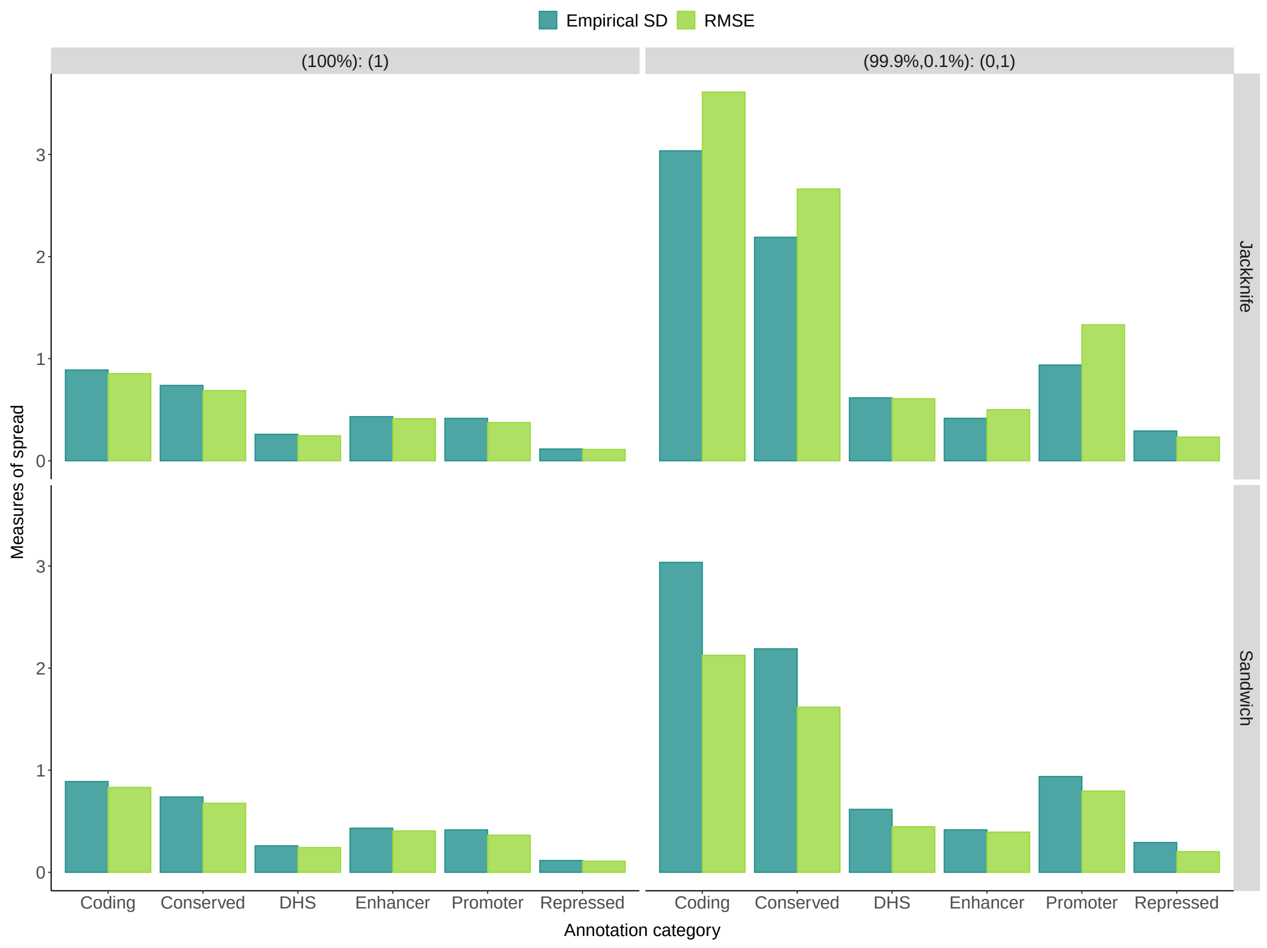


**Supplementary Figure 8. Standard error calibration using the jackknife vs. the sandwich estimator.** The column panel corresponds to different mixture components for generating the true causal effect sizes; the row panel corresponds to different SE estimators. Root mean square error (RMSE) is computed as the square root of the sum of the square SE divided by the number of experiments (100). Similarity between the empirical SD and the RMSE indicates good calibration.

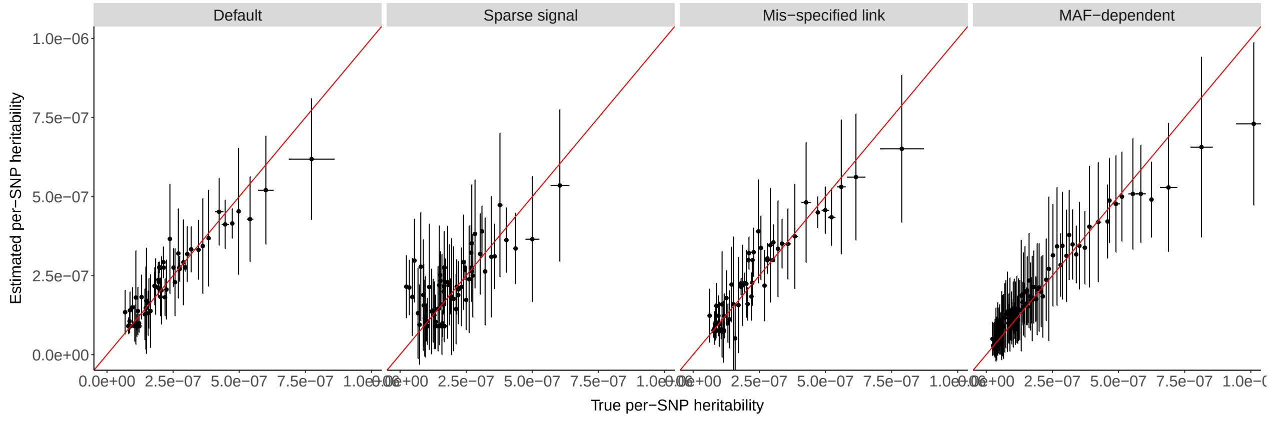


**Supplementary Figure 9. Calibration of per-SNP heritability under different model mis-specifications.** The points and the error bars on the x (y) axis correspond to the average and the SD of the true (estimated) per-SNP heritability. Colored in red is the 45 degree reference line. Alignment with the reference line indicates good calibration. The column panels represent different generative models or genetic architectures, same as those defined in **Figure** 1b of the main text.

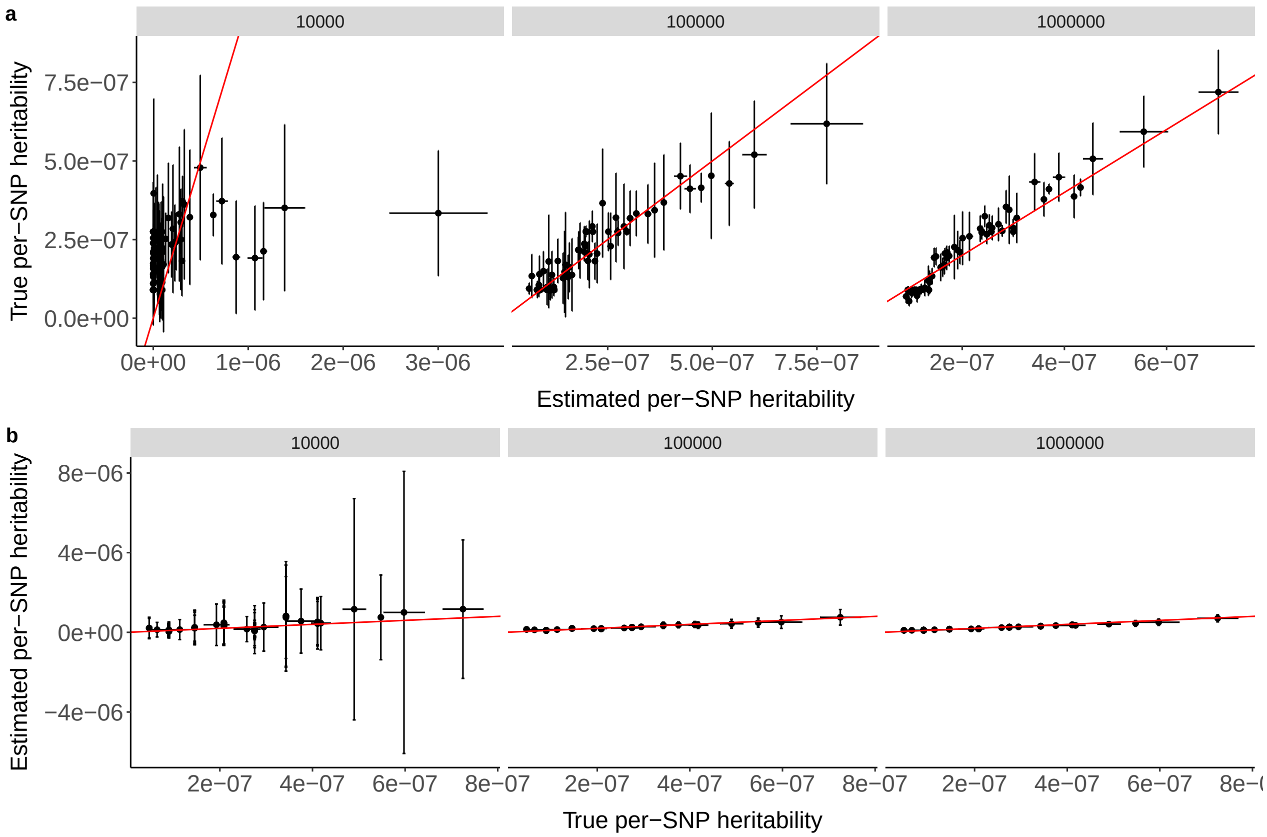


**Supplementary Figure 10.** Calibration of per-SNP heritability estimates from graphREML with different GWAS sample sizes. Summary statistics are directly simulated based on the LDGM precision matrices on chromosome 1 from the Europeans in 1000 Genome ($p = 513,012, n = 100,000$). SNPs are first ordered by the true (estimated) per-SNP genetic variances and then assigned to 100 equal-sized bins, based on the percentile values in the top (bottom) row. Panel names represent the GWAS sample size. Each point represents the average true (or estimated) SNP-heritability on the X-axis (or Y-axis), respectively. The error bars correspond to the SD of the true or estimated SNP-heritability within the bin. The red reference is the 45 degree line, $y = x$.

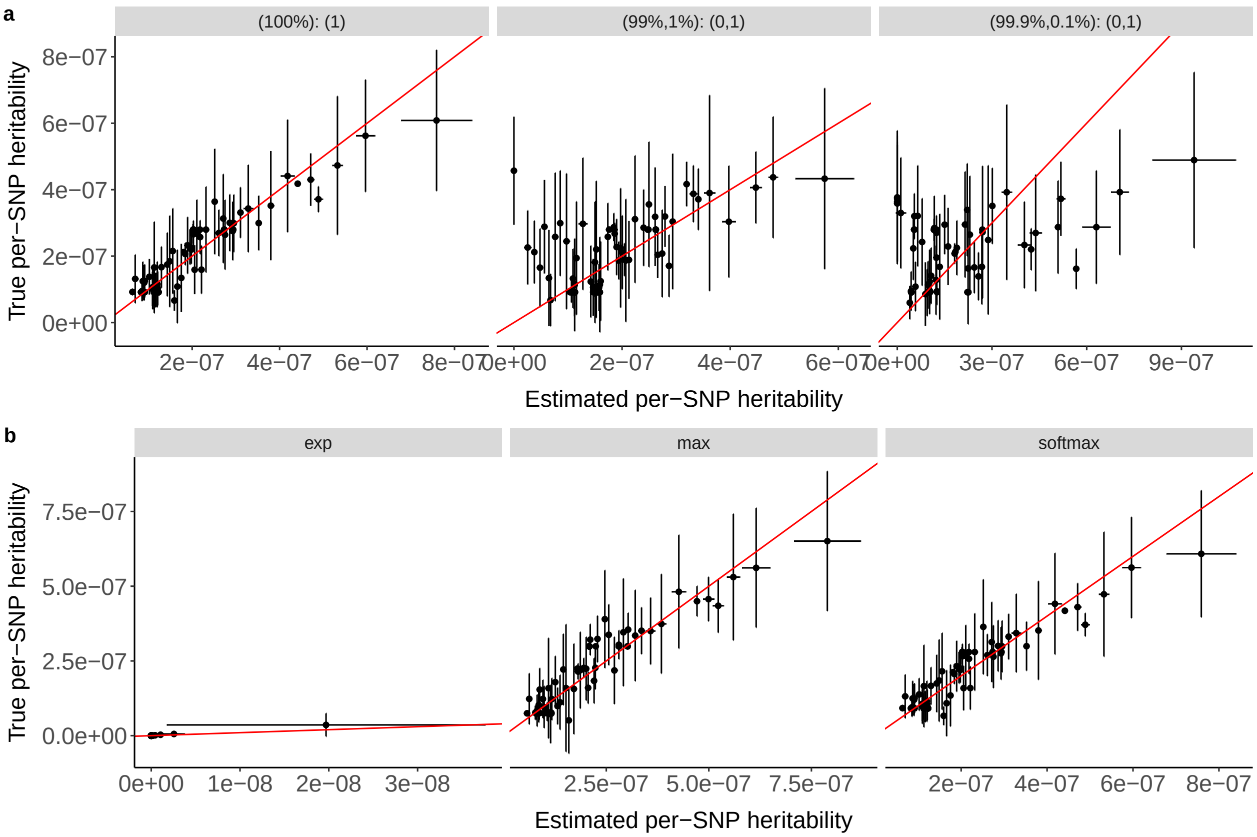


**Supplementary Figure 11.** Calibration of per-SNP heritability estimates from graphREML with different GWAS sample sizes. Summary statistics are directly simulated based on the LDGM precision matrices on chromosome 1 from the Europeans in 1000 Genome ($p = 513,012, n = 100,000$). SNPs are first ordered by the true (estimated) per-SNP genetic variances and then assigned to 100 equal-sized bins, based on the percentile values in the top (bottom) row. Panel names represent the GWAS sample size. Each point represents the average true (or estimated) SNP-heritability on the X-axis (or Y-axis), respectively. The error bars correspond to the SD of the true or estimated SNP-heritability within the bin. The red reference is the 45 degree line, $y = x$.

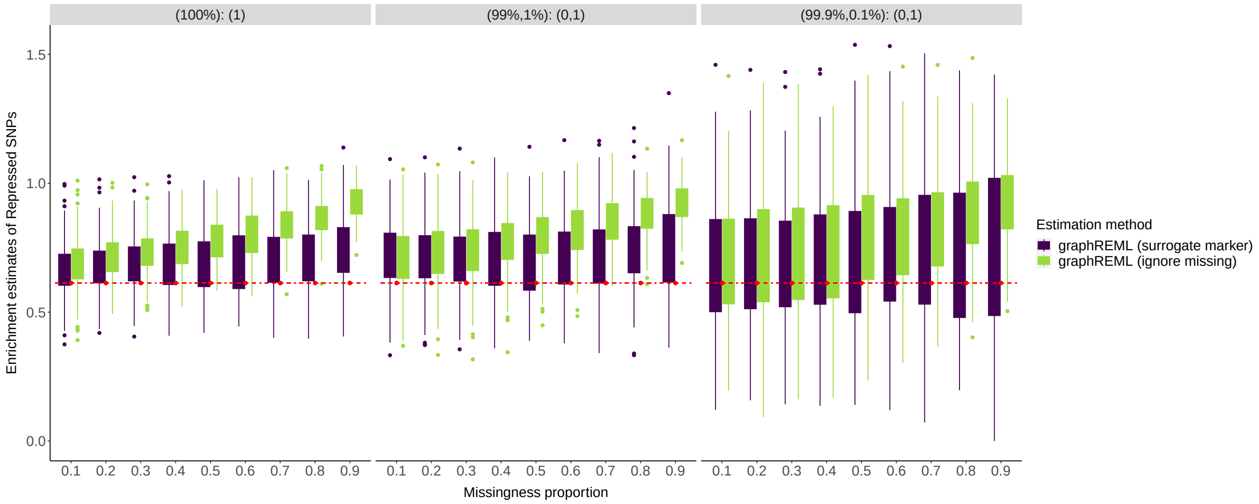


**Supplementary Figure 12. Impact of missingness on enrichment (depletion) estimates**. Summary statistics are directly simulated based on the LDGM precision matrices on chromosome 1 from the Europeans in 1000 Genome ($p = 513,012, n = 100,000$). The x-axis represents the proportion of SNPs in the LDGM that are missing in the summary statistics. The column panels represent different genetic architectures or mixture of causal effects, denoted as (component weights): (variance of the normal components). For example, (99%, 1%): (0,1) signifies that 99% of the SNPs are null and 1% of markers have their effect sizes drawn from $N(0,1)$. The red dashed line represents the true depletion value of the repressed SNPs in simulation.

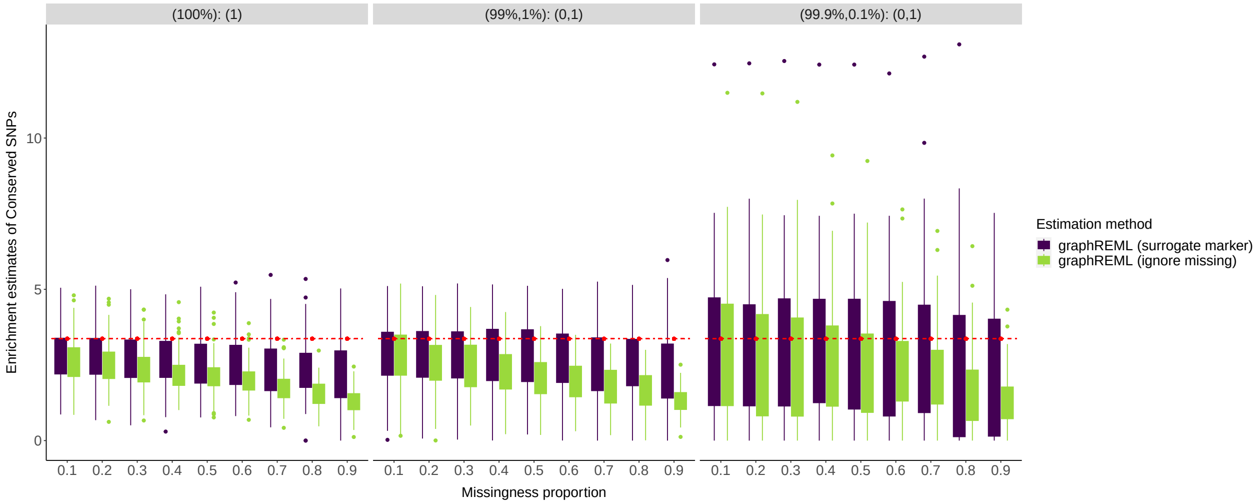


**Supplementary Figure 13. Impact of missingness on enrichment (depletion) estimates**. Summary statistics are directly simulated based on the LDGM precision matrices on chromosome 1 from the Europeans in 1000 Genome ($p = 513,012, n = 100,000$). The x-axis represents the proportion of SNPs in the LDGM that are missing in the summary statistics. The column panels represent different genetic architectures or mixture of causal effects, denoted as (component weights): (variance of the normal components). For example, (99%, 1%): (0,1) signifies that 99% of the SNPs are null and 1% of markers have their effect sizes drawn from $N(0,1)$. The red dashed line represents the true enrichment value of the conserved SNPs in simulation.

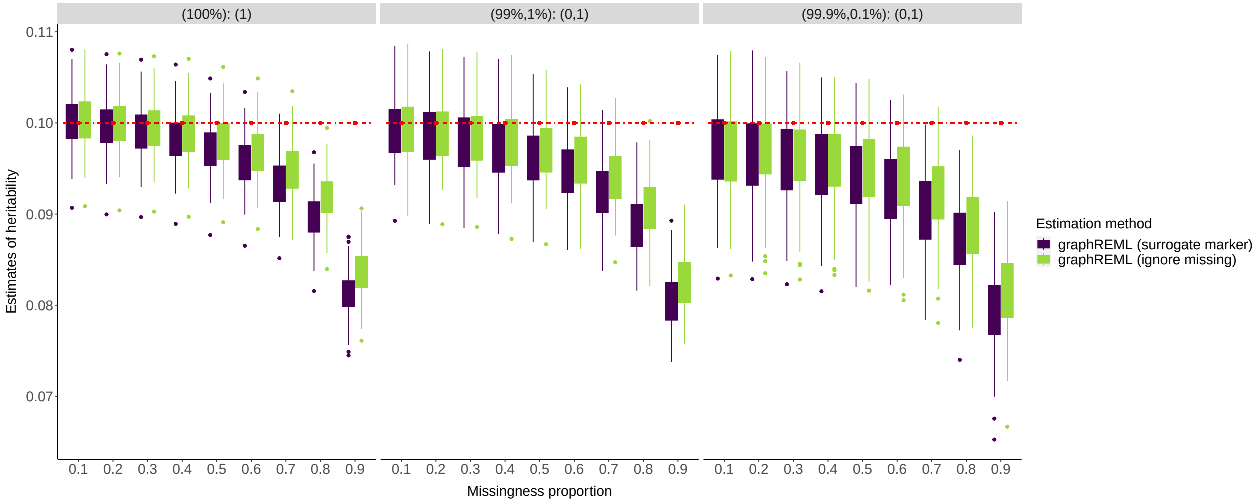


**Supplementary Figure 14. Impact of missingness on heritability estimates**. Summary statistics are directly simulated based on the LDGM precision matrices on chromosome 1 from the Europeans in 1000 Genome ($p = 513,012, n = 100,000$). The x-axis represents the proportion of SNPs in the LDGM that are missing in the summary statistics. The column panels represent different genetic architectures or mixture of causal effects, denoted as (component weights): (variance of the normal components). For example, (99%, 1%): (0,1) signifies that 99% of the SNPs are null and 1% of markers have their effect sizes drawn from $N(0,1)$. The red dashed line represents the true heritability in simulation.

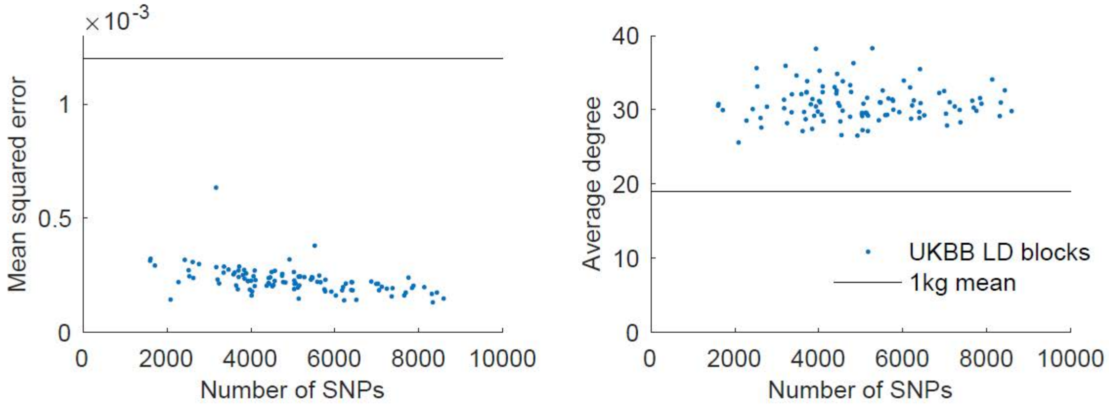


**Supplementary Figure 15.** **UK Biobank-derived LDGM precision matrices**, using 1000 Genomes (1000G) tree sequence-derived LDGMs from Nowbandganhi & Wohns et al.[2]. We computed new precision matrices from UK Biobank (UKBB)-derived LD matrices (N=460k self-reported white)[3]. We used v2 imputed array data and SNPs with MAF > 0.01 in 1000G European samples. To handle missingness in the UK Biobank data vs. 1000G Europeans, we (1) set missing values to the 1000 Genomes EUR allele frequency; (2) concatenated the 1,000 haploid European 1000 Genomes samples with the 460k diploid UKBB samples; (3) computed a correlation matrix from the concatenated genotypes; and (4) computed a sparse precision matrix from this correlation matrix and the ldgm software with default parameter settings. This approach avoids the need to re-create the LDGM in order to avoid missing variants; a naive approach of simply removing missing variants performed poorly. It is expected to have little effect on the correlation matrix (including the mean squared error) because the number of 1000G samples is so much smaller than the number of UKBB samples, and the number of missing SNPs was also small. (a) Mean squared error of inferred UKBB precision matrices vs. concatenated UKBB+1000G correlation matrices on chromosome 1, with a comparison to the average MSE between the 1000G EUR precision matrices vs. UKBB+1000G correlation matrices. Each datapoint is one LD block, and the number of SNPs per block is also listed. The reduction in MSE is expected due to increased sample size. (b) Average number of neighbors per SNP for UKBB precision matrices.

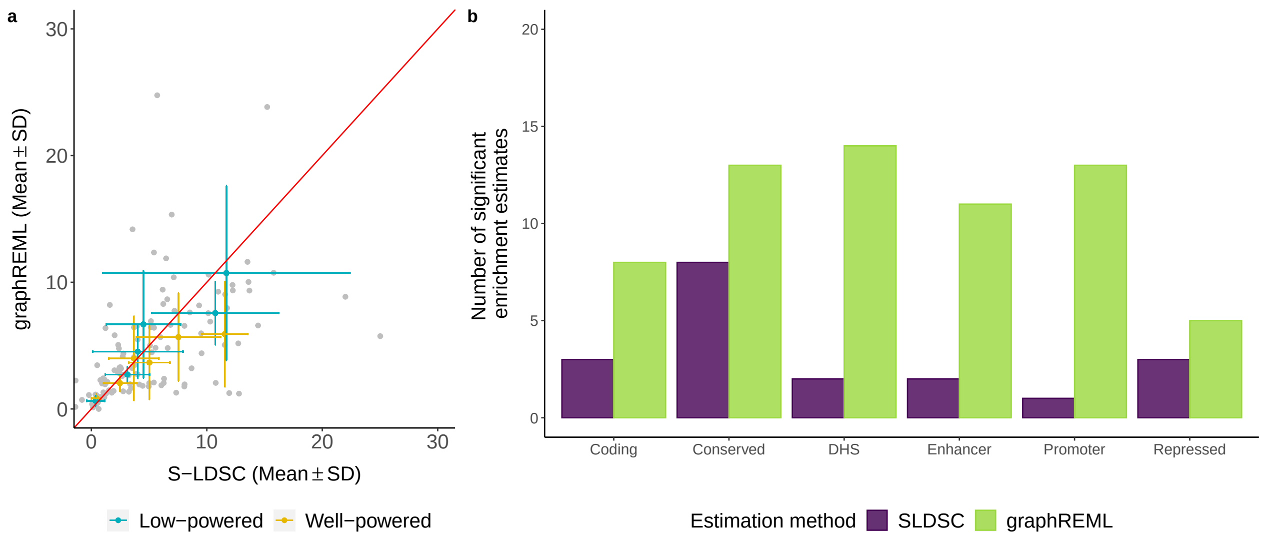


**Supplementary Figure 16.** Comparison of enrichment estimates from real trait analyses when both graphREML vs S-LDSC use out-of-sample LD. The association summary statistics are based on UK Biobank and the LDGMs are based on 1000 Genome.

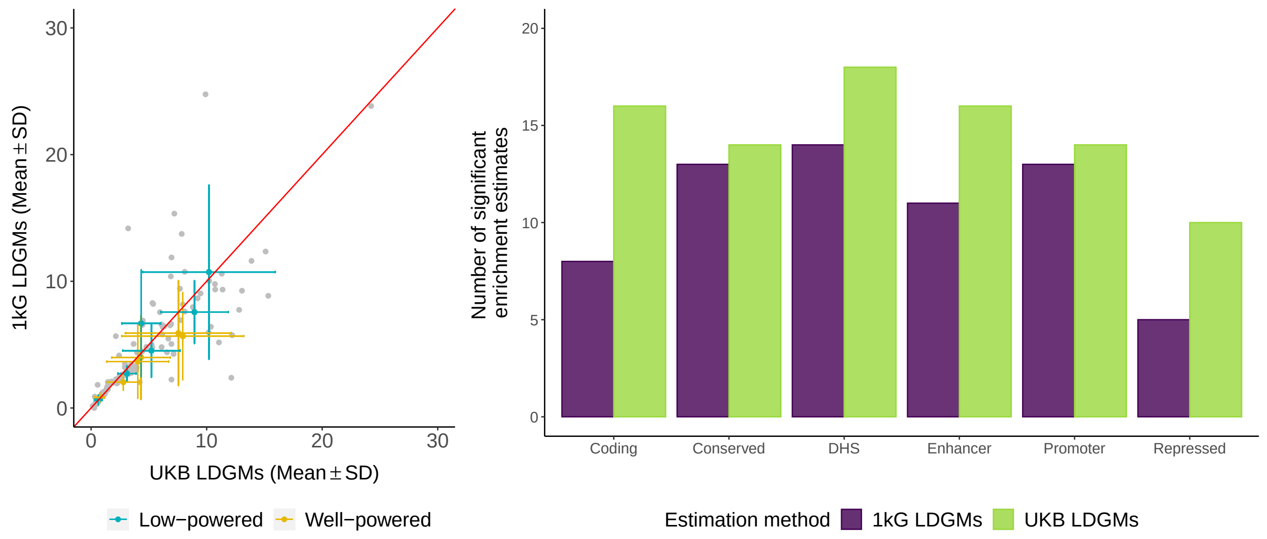


**Supplementary Figure 17.** Comparison of enrichment estimates from real trait analyses using in-sample (UKB) vs. out-of-sample (1kG) LDGMs. The set of phenotypes is the same as those used for method comparisons in **Figure** 3 of the main text. All estimates are based on graphREML.

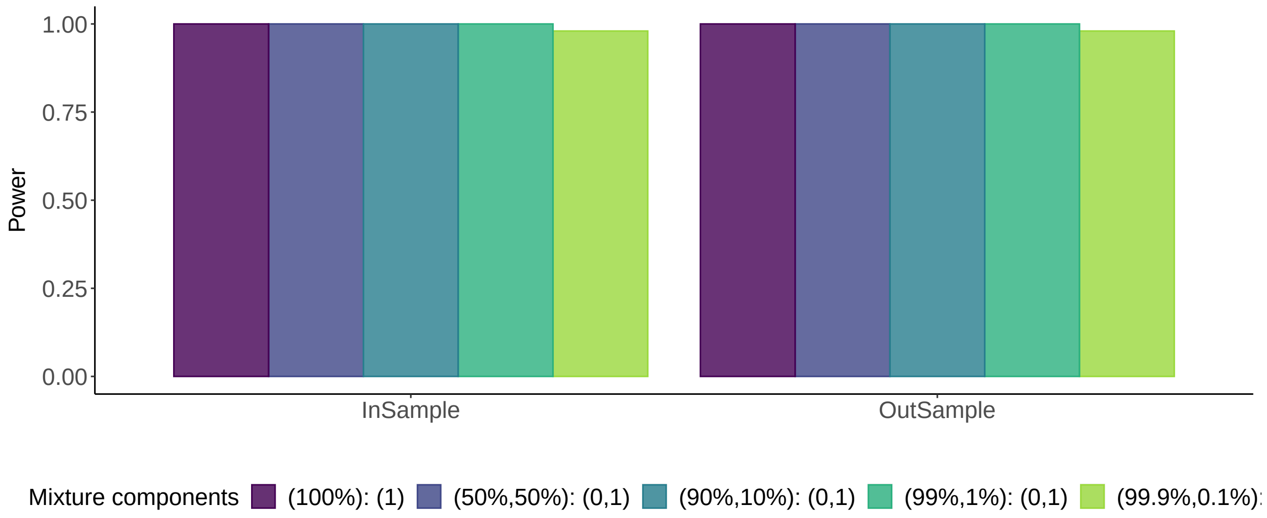


**Supplementary Figure 18. Power of the joint enrichment estimates.** Y-axis is the proportion of true non-nulls that have been correctly rejected or identified as significant. Genetic architectures are distinguished by the mixture components used to model the distribution of causal genetic effects, denoted as (component weights): (variance of the normal components). For example, (99%, 1%): (0,1) signifies that 99% of the SNPs are null and 1% of markers have their effect sizes drawn from N(0,1).

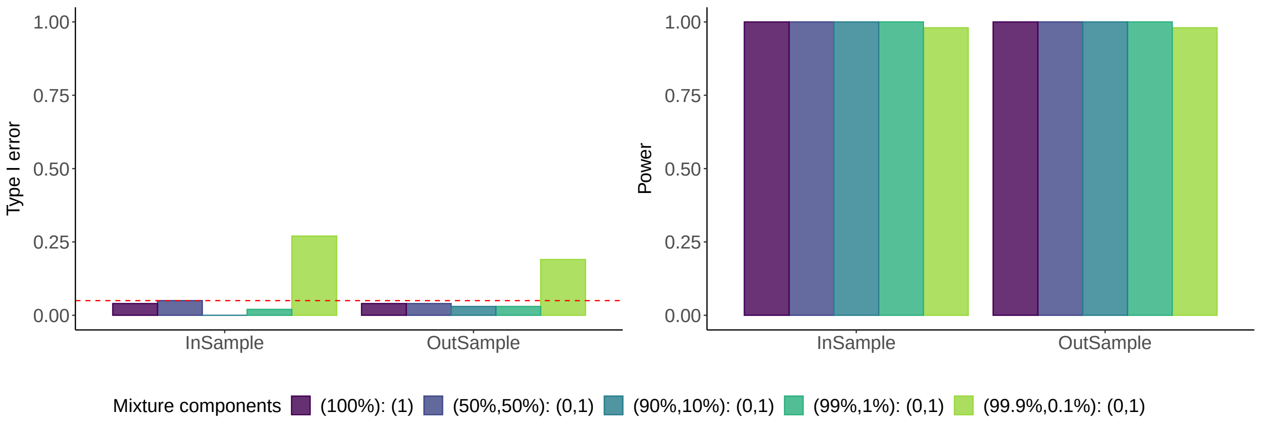


**Supplementary Figure 19.** The left panel is similar to **Figure** 1c in the main text; the right panel is similar to **Supplementary Figure** 17. The difference is that the null annotation is much smaller – coding region (1.8% of SNPs).

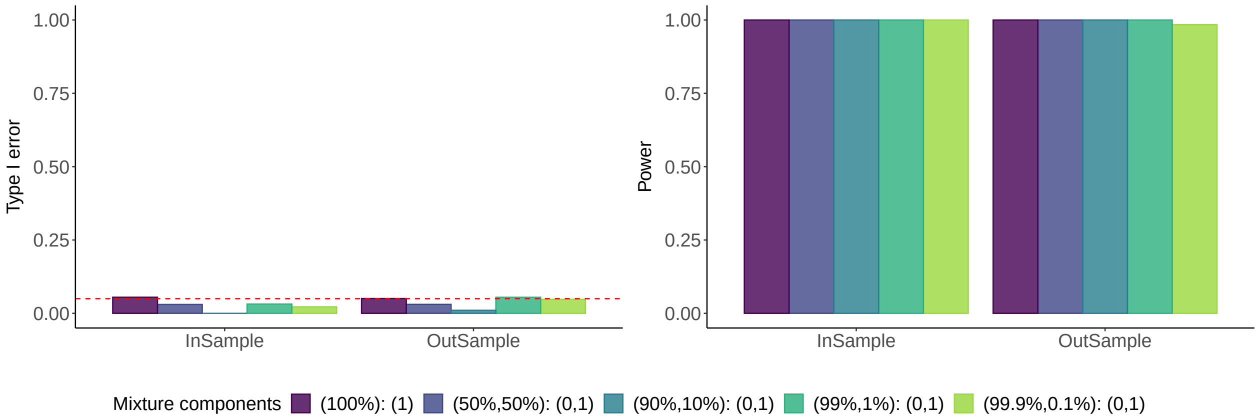


**Supplementary Figure 20.** The left panel is similar to **Figure** 1c in the main text; the right panel is similar to **Supplementary Figure** 17. The difference is that the sandwich estimator of SE is used instead of the jackknife estimator (**Online Methods**).

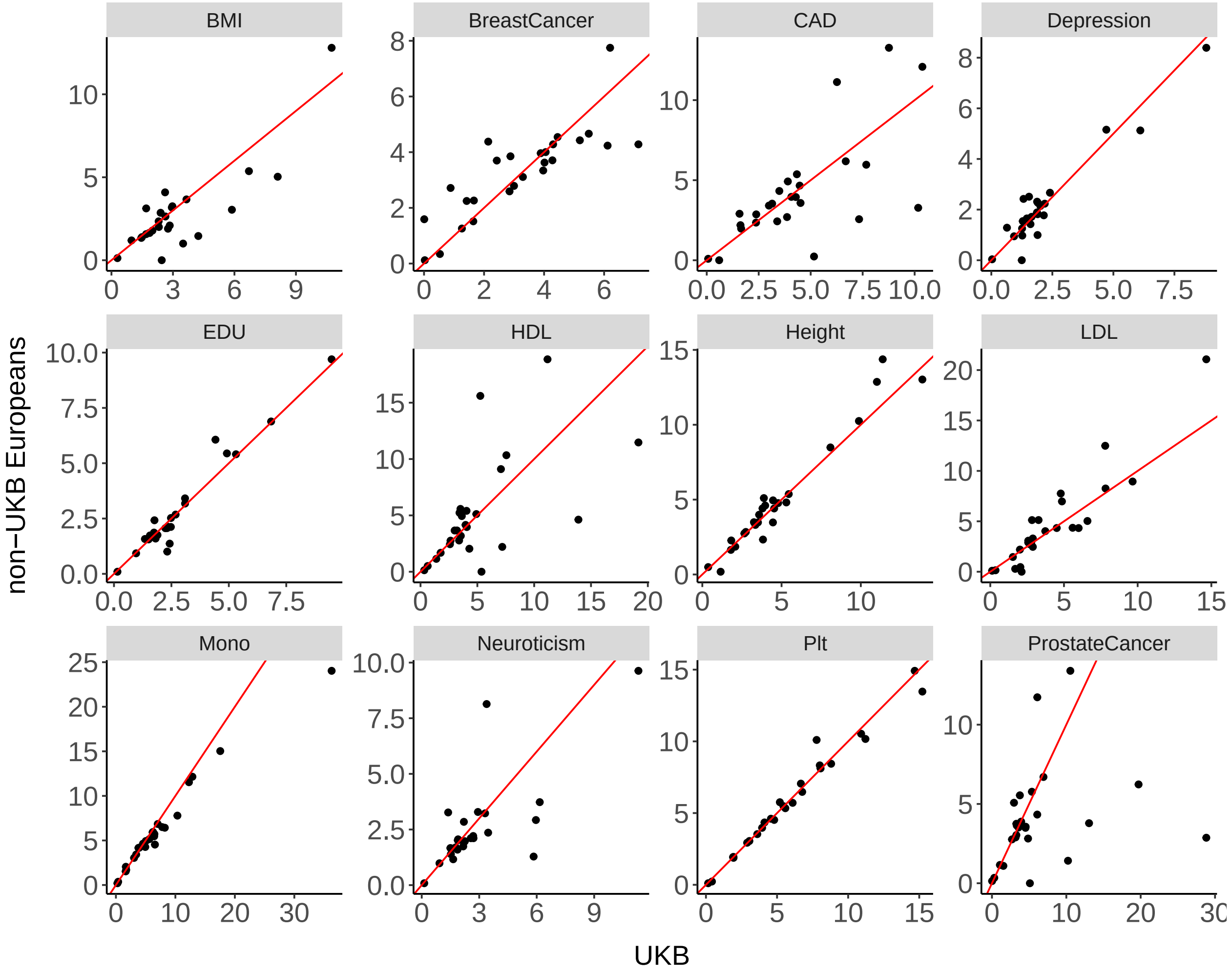


**Supplementary Figure 21.** Comparison of enrichment estimates when in-sample vs. out-of-sample LDGM precision matrices are used. Enrichment estimates based on the UKB GWAS are plotted on the x-axis; enrichment estimates based on the GWAS of non-UKB Europeans (*i.e.*, UKB is not the main component of the sample) are plotted on the y-axis. The red reference line is the 45 degree line. A summary of the summary statistics used in the comparison is provided in **Supplementary Table** 9. The numerical values of the estimates are reported in **Supplementary Table** 11.

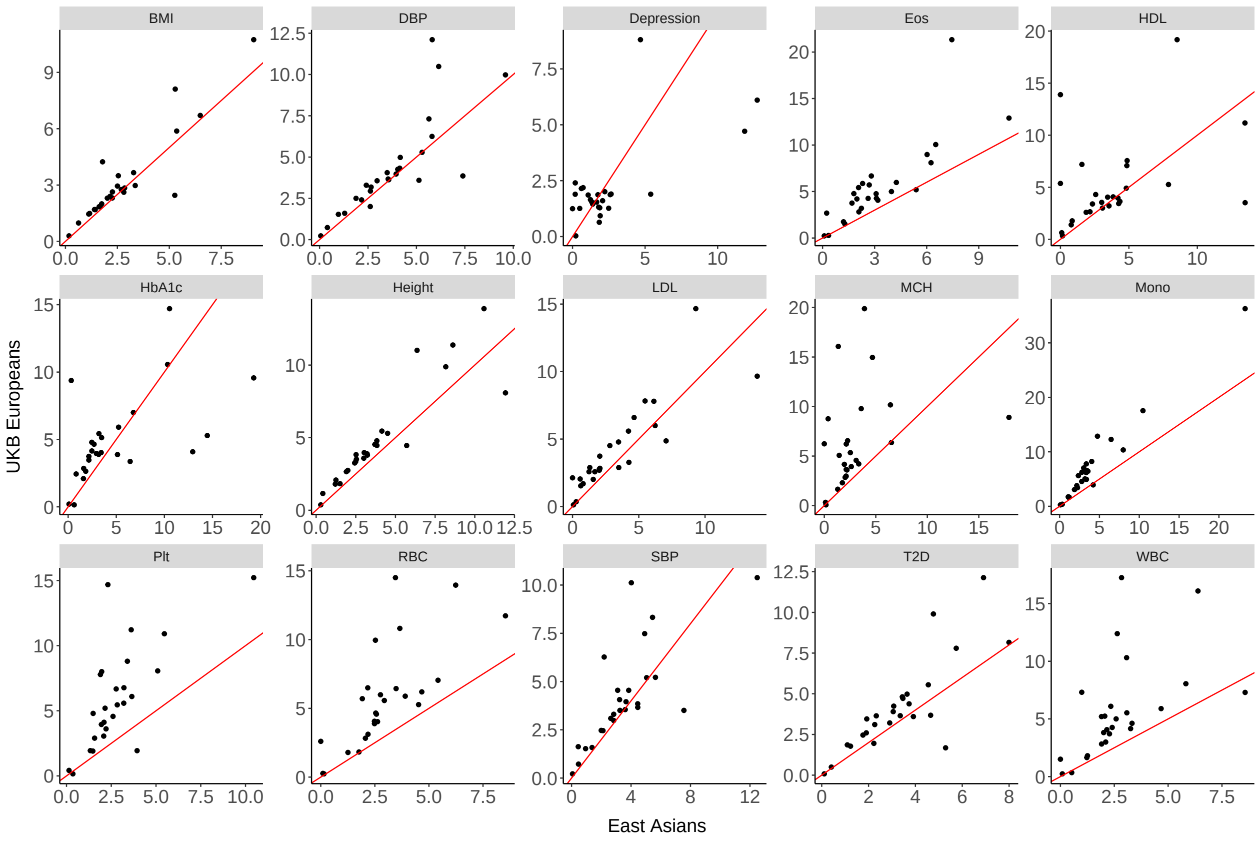


**Supplementary Figure 22.** Comparison of enrichment estimates between two ancestry groups. GWAS summary statistics and LDGM precision matrices inferred from matching ancestries are used to produce the enrichment estimates. LDGM precision matrices derived from the East Asian individuals in the 1000 Genome are used with the East Asian GWAS summary statistics. The red reference line is the 45 degree line. The numerical values of the estimates are reported in **Supplementary Table** 13.

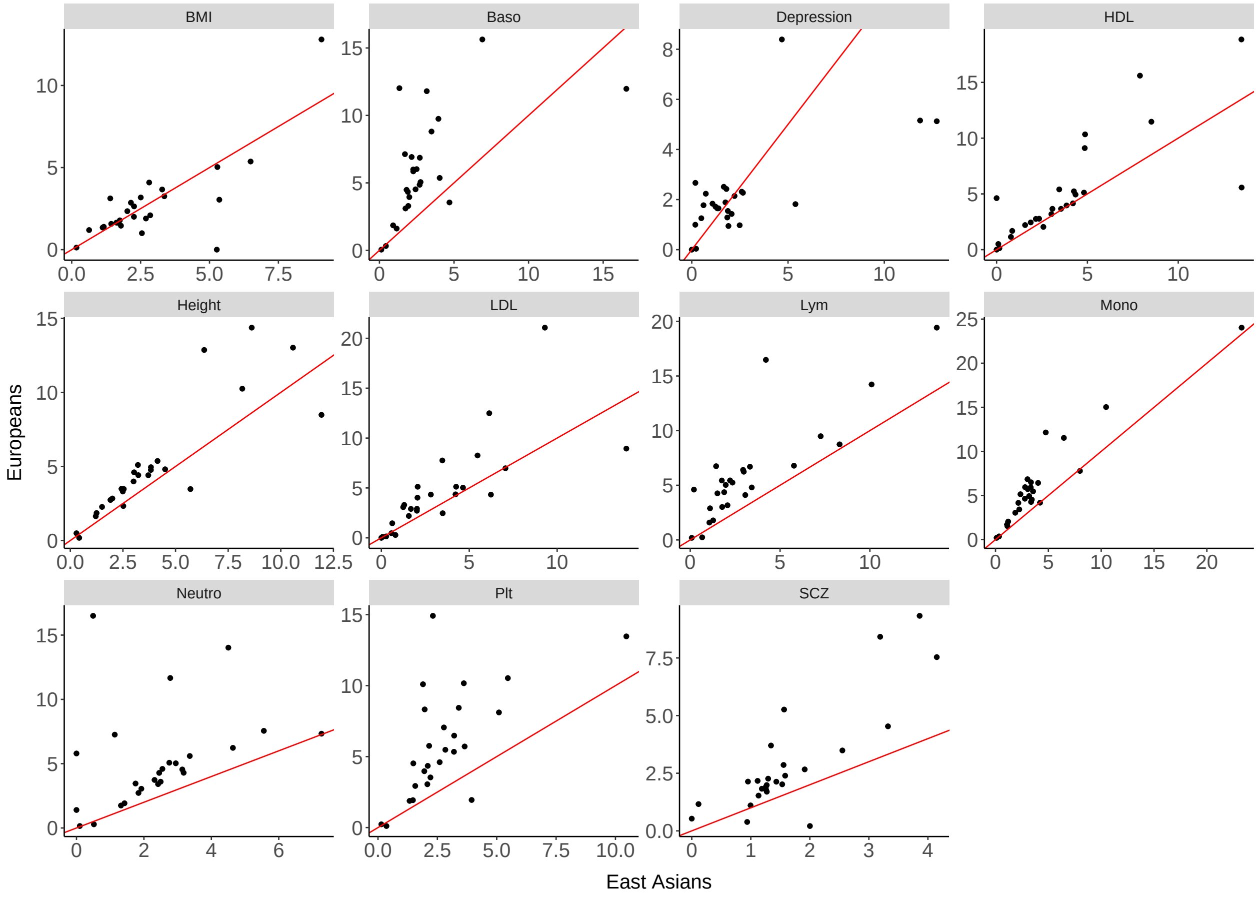


**Supplementary Figure 23.** Comparison of enrichment estimates between two ancestry groups. The difference to **Supplementary Figure** 19 is that plotted on the y-axis are the enrichment estimates based on GWAS summary statistics of European individuals (where UKB is not the major component of the sample). A summary of the summary statistics used in this comparison is provided in **Supplementary Table** 9-10. The numerical values of the estimates are reported in **Supplementary Table** 12.

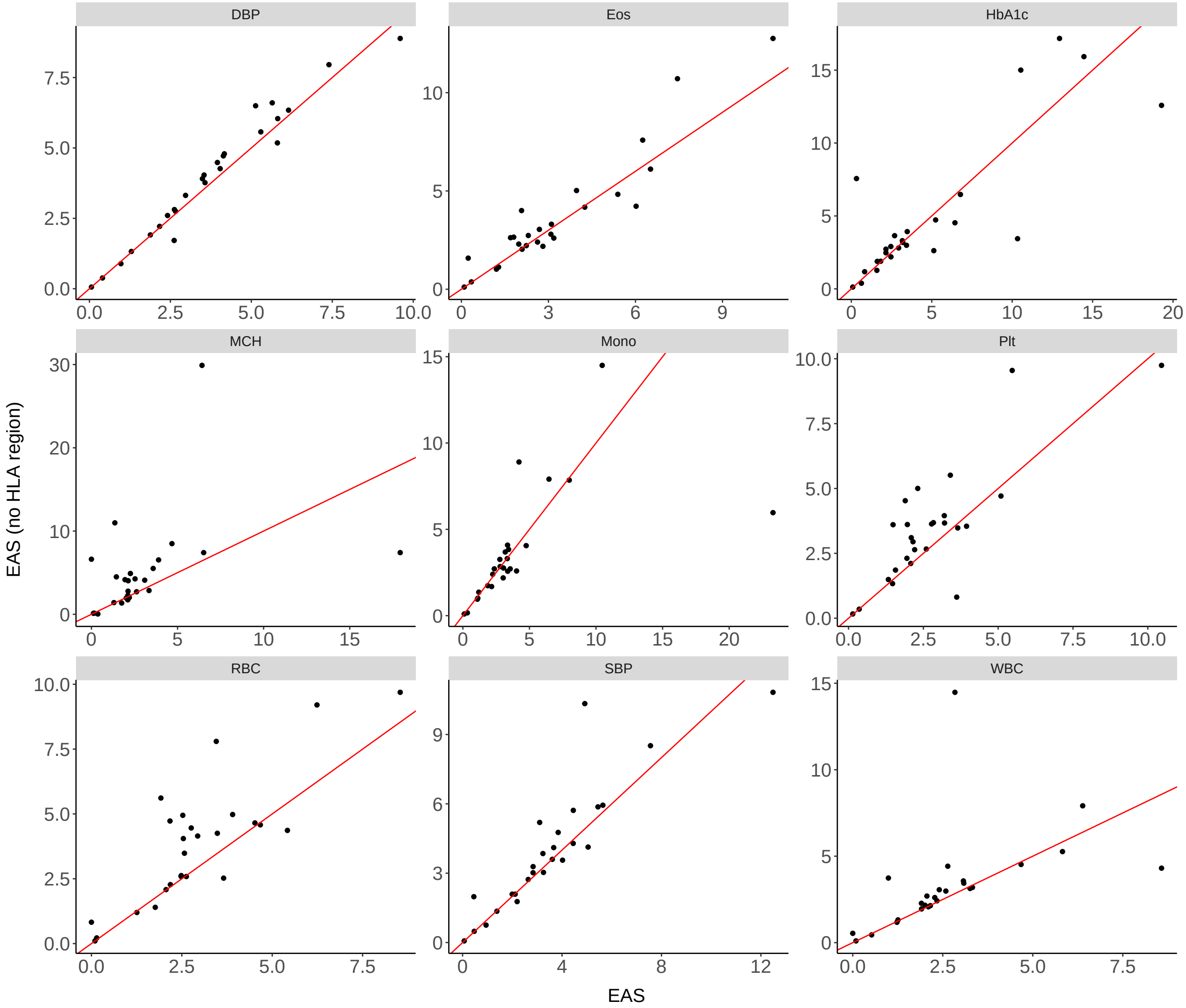


**Supplementary Figure 24.** Comparison of enrichment estimates with vs. without the variants in the HLA region included. The association statistics included in this analysis are blood or hematopoietic traits, for which we observed larger discrepancy in the enrichment estimates between Europeans and East Asians. LDGM precision matrices derived from the East Asian individuals in the 1000 Genome are used with the East Asian GWAS summary statistics. The red reference line is the 45 degree line.

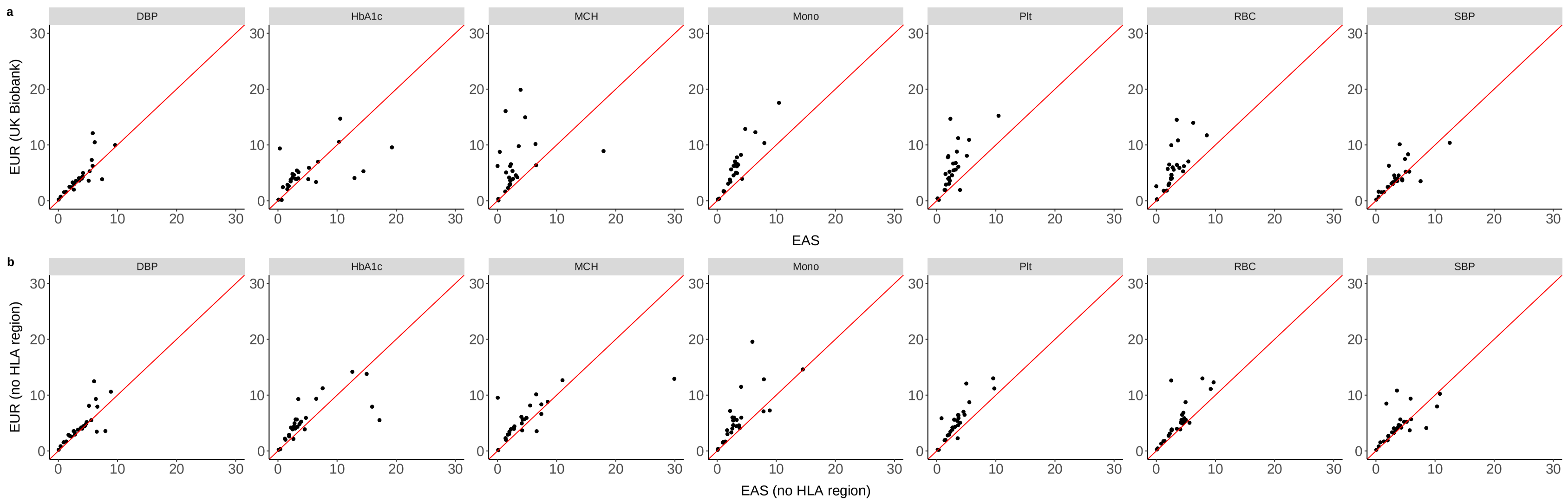


**Supplementary Figure 25. Impact of the HLA region on the cross-ancestry comparison of enrichment estimates.** Comparison of the enrichment estimates for a subset of blood and hematopoietic traits based on European samples vs. East Asian samples **a**. Variants in the HLA region are included in the enrichment estimates; same as **Supplementary Figure** 21, but reproduced here for contrast. **b**. Variants in the HLA region are excluded.

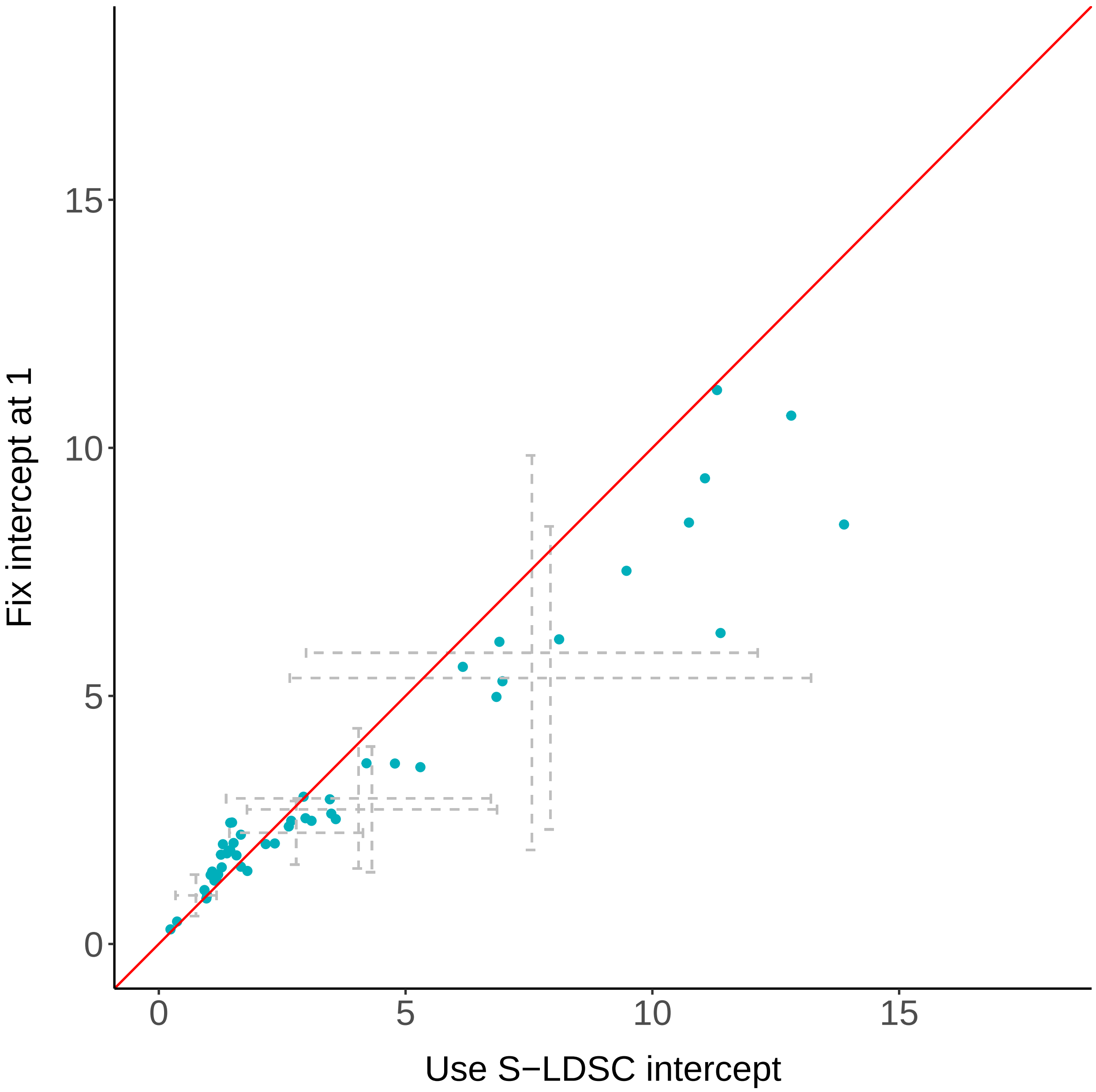


**Supplementary Figure 26. Impact of controlling for population stratification through S-LDSC.** Comparison of the graphREML results when we fix the intercept at 1 vs. at the S-LDSC estimated intercept. Blue dots are the enrichment estimates for the 18 validation traits (see descriptions of **Figure** 3 in the main analyses). Gray dotted lines are the meta-analyzed results for each annotation. Colored in red is the 45 degree reference line.
