## Supplementary Information for "Improved heritability partitioning and enrichment analyses using summary statistics with graphREML"

### Supplementary Notes

#### Statistical model

Let $\mathbf{y}$ be a length-$n$ vector that denotes the phenotypes of $n$ samples. Denote by $\mathbf{X} \in \mathbb{R}^{n \times p}$ the genotype matrix of $n$ individuals based on $p$ markers or SNPs. We standardize $\mathbf{X}$ and $\mathbf{y}$ such that the variance of the phenotype is 1 and the variance of each marker-specific genotype vector is 1. We use an additive genetic model for the phenotypes as $\mathbf{y} = \mathbf{X}\beta + \varepsilon$, where $\varepsilon$ is a length-$n$ vector distributed as $\varepsilon \sim N(0, \sigma_e^2 \mathbf{I}_n)$. We assume both $\mathbf{X}$ and $\beta$ are random, such that $\mathbf{\Sigma} \equiv \mathbb{E}(\mathbf{X}^\top \mathbf{X}/n)$ and $\mathbf{D} \equiv \mathbb{E}(\beta\beta^\top)$. The covariance of the effect sizes $\mathbf{D}$ is determined by our parameters of interest, such as partitioned heritability and enrichment of functional categories, which we denote by $\theta$. We use $\mathbf{D}(\theta)$ to emphasize the dependence of $\mathbf{D}$ on $\theta$.

Let $\mathbf{z} = \mathbf{X}^\top \mathbf{y}/\sqrt{n}$ be the vector of marginal association statistics from OLS. Then the distribution of $\mathbf{z}$, conditional on the true causal effect sizes can be shown as,

$$\mathbf{z}|\beta, \mathbf{X} \sim N(n^{-1/2}\mathbf{X}^\top \mathbf{X}\beta, n^{-1}\mathbf{X}^\top \mathbf{X}\sigma_e^2) \tag{1}$$

$$\mathbf{z}|\beta \sim N(n^{1/2}\mathbf{\Sigma}\beta, \mathbf{\Sigma}) \tag{2}$$

We emphasize that the distinction between the two distributions above lies in whether the genotype is conditioned on or assumed to be fixed. Our estimator is based on (2), which has been derived and used previously in the literature[1–3]. Suppose the causal effect sizes are drawn from a normal distribution, $\beta \sim N(0, \mathbf{D}(\theta))$. Integrating over $\beta$ leads to the marginal distribution,

$$\mathbf{z} \sim N(0, n\mathbf{\Sigma}\mathbf{D}(\theta)\mathbf{\Sigma} + \mathbf{\Sigma}), \tag{3}$$

and our estimator of $\theta$ comes from maximizing the likelihood function based on this marginal density. To make the algorithm computationally feasible, we work with the likelihood of $\tilde{\mathbf{z}} = \widehat{\mathbf{P}}\mathbf{z}$, where $\widehat{\mathbf{P}}$ is an estimate of $\mathbf{\Sigma}^{-1}$ based on LDGM, which is extremely sparse. It is easy to show that $\tilde{\mathbf{z}} \sim N(0, \mathbf{M}(\theta))$ where $\mathbf{M}(\theta) \equiv n\mathbf{D}(\theta) + \widehat{\mathbf{P}}$. We directly maximize the likelihood,

$$\ell(\theta) = -\frac{1}{2}\left\{log(2\pi) + log|\mathbf{M}(\theta)| + \tilde{\mathbf{z}}^\top \mathbf{M}(\theta)^{-\mathbf{1}}\tilde{\mathbf{z}}\right\}, \tag{4}$$

and the graphREML estimator is defined as $\widehat{\theta} = \arg\max_\theta \ell(\theta) = \arg\max_\theta \log \ell(\theta)$.

#### Estimation details

##### *Quasi-Newtonian algorithm*

We use a quasi-Newtonian algorithm for parameter estimation. This algorithm relies on efficient new subroutines, leveraging the sparsity of LDGM precision matrices to compute the likelihood function (4), together with its gradient and approximate Hessian. We iteratively update our estimate of the parameters as the following,

$$\theta^{(k+1)} = \theta^{(k)} - (\mathbf{H}^{(k)} + e\mathbf{I})^{-1}\nabla^{(k)},$$

where $\nabla^{(k)}$ and $\mathbf{H}^{(k)}$ are the gradient and the approximate Hessian of the likelihood function evaluated at the current estimate of the parameters $\theta^{(k)}$. $e$ is some small-valued number that is added to the diagonal of the Hessian matrix to prevent singularity in estimation. Let $\mathbf{M}(\theta^{(k)}) = n\mathbf{D}(\theta^{(k)}) + \widehat{\mathbf{P}}$. At each iteration, we first perform a Cholesky factorization of the matrix $\mathbf{M}(\theta^{(k)})$, which is feasible and computationally tractable due to the sparsity of $\widehat{\mathbf{P}}$. Denote by $\frac{\partial \mathbf{D_a}(\theta)}{\partial \theta_i}$ a diagonal matrix where the diagonal elements are the partial derivatives of the per-SNP heritability with respect to the parameters, $\left(\frac{\partial g_\theta(a_1)}{\partial \theta_i}, \ldots, \frac{\partial_\theta(a_p)}{\partial \theta_i}\right)$. We estimate the gradient using chain rule as the following,

$$\nabla_i^{(k+1)} = \frac{1}{2}n\left(\tilde{\mathbf{z}}^\top(\mathbf{M}^{(k)})^{-1}\frac{\partial \mathbf{D_a}(\theta)}{\partial \theta_i}(\mathbf{M}^{(k)})^{-1}\tilde{\mathbf{z}} - Tr\left(\frac{\partial \mathbf{D_a}(\theta)}{\partial \theta_i}(\mathbf{M}^{(k)})^{-1}\right)\right), \tag{5}$$

where we have used $\mathbf{M}^{(k)}$ to denote $\mathbf{M}(\theta^{(k)})$ for simplicity of notation, and $i$ indexes the parameters. Importantly, this equation is evaluated without computing $\mathbf{M}^{(k)}$ explicitly. In particular, the second term is evaluated by computing the sparse inverse subset of $\mathbf{M}^{(k)}$, as implemented in *suitesparse*[4]. We have four sparse matrix operations – matrix multiplication, division, log-determinant and matrix inverse. All of these functions were added to the LDGM package (see URL).

##### *Approximation of the Hessian matrix*

We use a second-order method to solve for the maximum likelihood estimators, which requires computing the Hessian matrix. The first derivative of (4) with respect to the $i$-th parameter at the $k$-th iteration is,

$$\nabla_i^{(k+1)} = \frac{1}{2}n\left(\tilde{\mathbf{z}}^\top(\mathbf{M}^{(k)})^{-1}\frac{\partial \mathbf{D}(\boldsymbol{\theta})}{\partial \boldsymbol{\theta}_i}(\mathbf{M}^{(k)})^{-1}\tilde{\mathbf{z}} - Tr\left(\frac{\partial \mathbf{D}(\boldsymbol{\theta})}{\partial \boldsymbol{\theta}_i}(\mathbf{M}^{(k)})^{-1}\right)\right). \tag{6}$$

The exact form of the second derivative of the log-likelihood with respect to the $i$-th and $l$-th parameters is,

$$\begin{aligned}\mathbf{H}_{il}^{(k+1)} = &\frac{1}{2}n\left[Tr\left(\frac{\partial^2 \mathbf{D}(\boldsymbol{\theta})}{\partial \theta_i \partial \theta_l}(\mathbf{M}^{(k)})^{-1}\right) - nTr\left(\frac{\partial \mathbf{D}(\boldsymbol{\theta})}{\partial \theta_i}(\mathbf{M}^{(k)})^{-1}\frac{\partial \mathbf{D}(\boldsymbol{\theta})}{\partial \theta_l}(\mathbf{M}^{(k)})^{-1}\right)\right] \\ &- \frac{1}{2}n\left[\tilde{\mathbf{z}}^\top\left((\mathbf{M}^{(k)})^{-1}\frac{\partial^2 \mathbf{D}(\boldsymbol{\theta})}{\partial \theta_i \partial \theta_l}(\mathbf{M}^{(k)})^{-1} - 2n(\mathbf{M}^{(k)})^{-1}\frac{\partial \mathbf{D}(\boldsymbol{\theta})}{\partial \theta_i}(\mathbf{M}^{(k)})^{-1}\frac{\partial \mathbf{D}(\boldsymbol{\theta})}{\partial \theta_l}(\mathbf{M}^{(k)})^{-1}\right)\tilde{\mathbf{z}}\right].\end{aligned} \tag{7}$$

The second line is easy to evaluate, but not the first line, which has two terms. Recall that $\tilde{\mathbf{z}} \sim N(0, \mathbf{M})$. Thus, we can use the trace trick for the expectation of a quadratic form and re-write the second term as the following,

$$-n^2 Tr\left(\frac{\partial \mathbf{D}(\boldsymbol{\theta})}{\partial \theta_i}(\mathbf{M}^{(k)})^{-1}\frac{\partial \mathbf{D}(\boldsymbol{\theta})}{\partial \theta_l}(\mathbf{M}^{(k)})^{-1}\right) = -n^2\mathbb{E}\left(\tilde{\mathbf{z}}^\top(\mathbf{M}^{(k)})^{-1}\frac{\partial \mathbf{D}(\boldsymbol{\theta})}{\partial \theta_i}(\mathbf{M}^{(k)})^{-1}\frac{\partial \mathbf{D}(\boldsymbol{\theta})}{\partial \theta_l}(\mathbf{M}^{(k)})^{-1}\tilde{\mathbf{z}}\right).$$

Similarly, we can approximate the first term in as the following,

$$nTr\left(\frac{\partial^2 \mathbf{D}(\boldsymbol{\theta})}{\partial \theta_i \partial \theta_l}(\mathbf{M}^{(k)})^{-1}\right) = n\mathbb{E}\left(\tilde{\mathbf{z}}^\top(\mathbf{M}^{(k)})^{-1}\frac{\partial^2 \mathbf{D}(\boldsymbol{\theta})}{\partial \theta_i \partial \theta_l}(\mathbf{M}^{(k)})^{-1}\tilde{\mathbf{z}}\right)$$

Next, we adopt the same trick as used in BOLT-REML[5], replacing the expected information with the observed information. With this approximation, the terms for the first and second line of equation (7) can be canceled out, which leads to the following,

$$\mathbf{H}_{il}^{(k+1)} \approx \frac{1}{2}n^2\tilde{\mathbf{z}}^\top\left((\mathbf{M}^{(k)})^{-1}\frac{\partial \mathbf{D}(\boldsymbol{\theta})}{\partial \theta_i}(\mathbf{M}^{(k)})^{-1}\frac{\partial \mathbf{D}(\boldsymbol{\theta})}{\partial \theta_l}(\mathbf{M}^{(k)})^{-1}\right)\tilde{\mathbf{z}}, \tag{8}$$

##### ***Trust-region algorithm***

We use the trust-region algorithm to control the step size of each update in a principled way, and employ an adaptive bound on the maximum change at each iteration (**Algorithm** 1). This allows us to balance between convergence speed and robustness of the updates. We referenced the trust region iterative optimization in Loh et al.[6] in developing our own algorithm, but modified several aspects of the procedure for graphREML. The main differences are that: 1) we compute the actual log-likelihood for the decision rule to accept or reject the step size; 2) we do not impose the constraint related to parameter domain, because our link function automatically leads to non-negative per-SNP heritability and thus a valid covariance matrix (for the effect sizes); 3) we do not explicitly *optimize* the step size by maximizing the predicted change in log likelihood according to the local quadratic model since this optimization can incur further computational cost; instead, we simply use the closed-form step size similar to that used in Newton Raphson.

A key quantity that is central to both step size acceptance and trust region radius update is the ratio between the actual and the predicted change of log likelihood, $\rho = \Delta_{actual}/\Delta_{pred}$. We use the hyperparameter values recommended by Gould et al.[7] to set up the lower and upper bounds, denoted by $\underline{\rho}$ and $\bar{\rho}$ respectively. These parameters, along with the radius change rate $\mu_{TR}$, determine the updating of the trust region radius adjustment.

The trust region algorithm is embedded within the Newton Raphson algorithm, such that each update involves a step size adaptation. Therefore, each trust-region algorithm takes the current values of the parameters as input and output the updated parameter values, along with the selected step size. Note that the trust region radius is also adjustable, which is passed down from one iteration to the next. In addition, we adopt the safeguard procedure proposed in Loh et al.[6], rejecting step sizes that lead to an updated gradient whose norm is more than double the norm of the gradient evaluated at the current parameter values.

##### ***Convergence Criterion***

We stop the Newton Raphson algorithm either when the maximum number of updates have reached or if the convergence criterion is triggered. We use a rather stringent threshold to determine convergence:

**Algorithm 1: Trust region algorithm for step size selection**

**Preset** : $\mu_{TR} = 10$; $\underline{\rho} = 10^{-4}$; $\bar{\rho} = 0.99$; $K = 20$
**Input** : $\theta^k$
**Init** : $\lambda_{TR} = 10^{-3}$; $k = 0$; accept=0

**while** *$k < K$ and accept is 0* **do**
  $k \longleftarrow k + 1$;
  $\widetilde{\mathbf{H}} \longleftarrow \mathbf{H}(\theta^k) + 0.01 \cdot \lambda_{TR}(\mathbf{I} \odot \mathbf{H}(\theta^k))$;
  $\mathbf{s} \longleftarrow \widetilde{\mathbf{H}}^{-1}\nabla(\theta^k)$ ; /* Compute the proposed step size */
  $\widetilde{\theta} \longleftarrow \theta^k - \mathbf{s}$;
  $\Delta_{actual} \longleftarrow \ell(\theta^k) - \ell(\theta^k)$;
  $\Delta_{pred} \longleftarrow \mathbf{s}^\top \nabla(\theta^k) - \frac{1}{2}\mathbf{s}^\top \mathbf{H}(\theta^k)\mathbf{s}$
  $\rho = \Delta_{actual}/\Delta_{pred}$ ; /* Evaluate the step size */
  **if** $\rho > \underline{\rho}$ **then**
    $\theta^{k+1} \longleftarrow \widetilde{\theta}$; accept = 1;
  **else**
    $\theta^{k+1} \longleftarrow \widetilde{\theta^k}$ ; /* Accept or reject the step size */
  **end**
  **if** $\rho < \underline{\rho}$ **then**
    $\lambda_{TR} = \lambda_{TR} \cdot \mu_{TR}$;
  **else**
    **if** $\rho > \bar{\rho}$ **then**
      $\lambda_{TR} = \lambda_{TR}/\mu_{TR}$ ; /* Update the trust region parameters */
    **end**
  **end**
**end**

**Output** : $\theta^{k+1}$; $\mathbf{s}$, $\lambda_{TR}$;

we set the maximum number of iterations to 50, and declares convergence of the algorithm when the change of log-likelihood averaged over three consecutive iterations is less than $10^{-3}$. The sensitivity of the estimation results with respect to these hyperparameters can vary depending on the dataset and the size of the problem (*i.e.*, number of parameters). These parameters can be adjusted easily if needed.

#### Modeling the per-SNP heritability

S-LDSC assumes an unrealistic linear relationship between the heritability of a SNP and its annotations, leading for example to negative per-SNP heritability estimates. In contrast, graphREML can fit essentially any heritability model, incorporating a flexible link function to map between the annotations of a SNP to its heritability. We assume that $\mathbf{D}$ is a diagonal matrix with non-negative diagonal elements that represent the per-SNP heritability of SNPs in the model. In other words, we assume that the covariance of $\tilde{\mathbf{z}}$ has the form,

$$\mathbf{M}(\theta) = n \cdot diag(\sigma_1^2, ..., \sigma_p^2) + \widehat{\mathbf{P}}, \quad \text{with} \quad \sigma_j^2 = g_\theta(\mathbf{a}_j), \tag{9}$$

where $\mathbf{a}_j$ denotes the vector of annotation values for SNP $j$, and $\theta$ denotes the conditional enrichment coefficients. $g_\theta$ is a non-negative scalar-valued link function that we choose for estimation. graphREML by default uses the softmax link function, $g(\mathbf{a}_j) = \log(1 + \exp(\mathbf{a}_j^\top \theta))$, but a more general form of $\mathbf{D}$ and other options for the link function are possible.

Notably, the existing methods can be viewed as special cases of (9), in terms of their assumptions about the covariance structure of the causal effect sizes. For example, S-LDSC assumes $\sigma_j^2 = \mathbf{a}_j^\top \theta$, where $\theta$ is a vector of coefficients that determine the partitioned heritability and enrichment[8,9]. SumHer varies from S-LDSC in modeling $\sigma_j^2$ as $\mathbf{a}_j^\top \theta q_j / Q$, where $q_j$ is a weight that explicitly accounts for the frequency-dependent and LD-dependent architecture, and $Q$ is the normalizing constant such that $Q = \sum_{j=1}^{p} q_j$[10]. It is evident from the modeling of $\sigma_j^2$ that the per-SNP heritability estimates from S-LDSC or SumHer may be negative and thus invalid. In contrast, graphREML is guaranteed to produce valid non-negative per-SNP heritability estimates, as long as a non-negative link $g(\cdot)$ is used.

##### *Numerical overflow*

One implementation detail associated with the softmax link function is that we observed an numerical overflow issue when applying the softmax function. To avoid this problem, we rewrote the link as the following:

$$
\begin{aligned}
g_\theta(\mathbf{a}_j) &= \log(1+\exp(\mathbf{a}_j^\top \theta)) \\
&= log\left(1+\exp\left(\mathbf{a}_j[-]\cdot\theta[-]+\mathbf{a}_j[+]\cdot\theta[+]\right)\right) \\
&= log\left(\frac{\exp\left(\mathbf{a}_j[-]\cdot\theta[-]\right)+\exp\left(-\mathbf{a}_j[+]\cdot\theta[+]\right)}{\exp\left(-\mathbf{a}_j[+]\cdot\theta[+]\right)}\right) \\
&= \mathbf{a}_j[+]\cdot\theta[+] + log\left[\exp\left(\mathbf{a}_j[-]\cdot\theta[-]\right)+\exp\left(-\mathbf{a}_j[+]\cdot\theta[+]\right)\right].
\end{aligned}
\tag{10}
$$

where $[+]$ and $[-]$ indicate the subset of the parameters which are positive and negative, respectively. We implemented a version of the softmax function using equation (10), which guards against numerical overflow issues. Analogously, we implemented a more robust version of the link derivative as the following,

$$
g'_\theta(\mathbf{a}_j) = \frac{\mathbf{a}_j[-]\cdot\exp\left(\mathbf{a}_j[-]\cdot\theta[-]\right)}{1+\exp\left(\mathbf{a}_j[-]\cdot\theta[-]\right)} + \frac{\mathbf{a}_j[+]}{1+\exp\left(-\mathbf{a}_j[+]\cdot\theta[+]\right)}. \tag{11}
$$

We scale the link and the link gradient functions by one over the number of SNPs. This ensures that any non-linear relationships between the SNP's annotation values and its heritability can be properly captured (the softmax function becomes effectively linear when $\mathbf{a}_j^\top\theta$ is very large).

##### ***Differentiability***

We note that it is important for the link function to be differentiable everywhere, because our estimation algorithm, the gradient of the likelihood function in particular, entails the first derivative of the link function. As we will discuss below, performing a score test for the conditional enrichment of an annotation requires taking the second derivative of the link function. For our default link function – the softmax – we have $\frac{\partial g(\mathbf{a}_j)}{\partial \theta_k} = \frac{\mathbf{a}_{j,k}\exp(\mathbf{a}_j^\top\theta)}{1+\exp(\mathbf{a}_j^\top\theta)}$, and $\frac{\partial^2 g(\mathbf{a}_j)}{\partial \theta_k \theta_l} = \frac{\mathbf{a}_{jk}\mathbf{a}_{jl}\exp(\mathbf{a}_j^\top\theta)}{(1+\exp(\mathbf{a}_j^\top\theta))^2}$. Other smooth candidate link functions include the logistic function, $g(\mathbf{a}_j) = \frac{1}{1+\exp(-\mathbf{a}_j^\top\theta)}$.

#### Score test for inference on the joint enrichment

Here we provide more details on the score test procedure we use for inference on the joint enrichment of a new annotation, conditional on a set of baseline annotations. The key observation that enables us to derive this test is that the score contributed from a given SNP can be re-written via chain rule into two parts – one part is the derivative of the likelihood with respect to the per-SNP heritability; the other part is the derivative of per-SNP heritability with respect to the parameters. In particular, when evaluated at the null, the score involves the new annotation values only through the second part.

Let $\hat{\theta} = (\hat{\theta}_1, \hat{\theta}_2, ..., \hat{\theta}_K)$ denote the set of $K$ parameter estimates in the baseline model. Let $\theta^* = (\hat{\theta}, 0)$ denote the parameters fitted under the null *i.e.*, with baseline annotations included and constraining the conditional enrichment of the new $K+1$-th annotation to be zero. The score contributed from SNP $j$ can be written as the following,

$$
\begin{aligned}
U_{j,K+1}(\theta^*) &= \frac{\partial \ell}{\partial \sigma_j^2} \cdot \frac{\partial \sigma_j^2}{\partial \theta_{K+1}} \bigg|_{\theta=\theta^*} \\
&= \mathbf{a}_{j,K+1} \left[ g'(\mathbf{a}_j^\top \theta) \frac{\partial \ell}{\partial g(\mathbf{a}_j^\top \theta)} \bigg|_{\theta=\theta^*} \right]_j && \text{(chain rule)} \\
&= \mathbf{a}_{j,K+1} \underbrace{\left[ g'(\mathbf{a}_j^\top \theta) \frac{\partial \ell}{\partial g(\mathbf{a}_j^\top \theta)} \bigg|_{\theta=\hat{\theta}} \right]_j}_{\text{SNP-specific gradient obtained from the null fit}} && (\theta^*_{K+1} = 0 \text{ and thus } \mathbf{a}_j^\top \theta^* = \mathbf{a}_j^\top \hat{\theta}) \\
&= \mathbf{a}_{j,K+1} \nabla_j(\hat{\theta}) && (12)
\end{aligned}
$$

Re-writting the score as equation (12) enables us to separate out the part that solely relies on the null fit and the part that entails the new annotation, which is the basis for the score test we developed.

The procedure we propose is as follows:

1. Fit graphREML under the null model (with $K$ baseline annotations) to obtain the SNP-specific gradients, $\nabla_j(\hat{\theta})$ for $j = 1, 2, ..., p$.

2. For any new annotation $\mathbf{a}_{.K+1}$, construct the following statistics,

$$S_{K+1} = \frac{U_{K+1}(\theta^*)^2}{Var(U_{K+1}(\theta^*))} \tag{13}$$

where $U_{K+1}(\theta^*) = \sum_j \mathbf{a}_{j,K+1} \nabla_j(\hat{\theta})$ is the score aggregated from all markers.

3. Compare the score statistics against the null distribution $S_{K+1} \overset{H_0}{\sim} \chi^2(1)$ to compute the p-value for the enrichment of this new annotation.

##### *Test of multiple new annotations*

More generally, we can adopt a similar procedure to test the significance of multiple new annotations at the same time, conditional on the baseline annotations. To jointly test the significance of $L$ new annotations conditional on the $K$ baseline annotations, we can construct the following test statistics,

$$S = U(\theta^*)^\top Cov(U(\theta^*)) U(\theta^*) \tag{14}$$

where $U(\theta^*) = [U_{K+1}(\theta^*), ... U_{K+L}(\theta^*)]^\top$. Under the null, $S \overset{H_0}{\sim} \chi^2(L)$, with which we can compute the p-value for the joint enrichment of these $L$ annotations.

##### *Jackknife covariance estimator*

We adopt a similar procedure as that used in the Wald test to derive a jackknife estimator of the SE. We take advantage of the LD block structure to calculate the empirical covariance of the scores as the plug-in variance for the score statistic, First, we compute a set of leave-one-LD-block scores,

$$J^b_{K+1}(\theta^*) = U_{K+1}(\theta^*) - \sum_{j \in b} \mathbf{a}_{j,K+1} \nabla_j(\hat{\theta}), \quad \forall b = 1, 2, ... B$$

Then, we use the empirical distribution of the jackknife scores to construct the score test,

$$\frac{1}{B-2} \frac{\left(\sum_b J^b_{K+1}(\theta^*)/B\right)^2}{Var(J^b_{K+1}(\theta^*))} \overset{H_0}{\sim} \chi^2(1).$$

##### *Accounting for the uncertainty in $\hat{\theta}$*

We note that $\hat{\theta}$ is not perfectly estimated, which can affect the score test in two ways – one is that the estimates upon termination of the Newton updates may not be the actual solution to the score equations or the maximizer of the likelihood; the other is that the estimation noise needs to be accounted for through the plug-in variance estimator of the score statistic.

First, we need to account for the fact that the likelihood function evaluated at $\hat{\theta}$ is not the actual maximum even though it may be sufficiently close, because we stop the Newton updates using a pre-specified convergence criterion. Denote by $\tilde{\theta}$ the true solutions to the score equations under the null. Then by definition,

$$U_k(\theta)|_{\theta=\tilde{\theta}} = \sum_{j\in[p]} \mathbf{a}_{j,k}\nabla_j(\tilde{\theta}) = 0, \forall k = 1,2,...,K \tag{15}$$

which is equivalent to $\mathbf{a}_k \perp \nabla(\tilde{\theta})$, for all $k = 1,2,...,K$. In practice, the estimate we obtain from the null fit $\hat{\theta}$ may be arbitrarily close but is not exactly $\tilde{\theta}$. We adjust the SNP-specific gradients by projecting $\nabla(\hat{\theta})$ onto the null space expanded by the baseline annotation matrix $\mathbf{A}$,

$$\nabla(\tilde{\theta}) \approx (\mathbf{I} - \mathbf{A}(\mathbf{A}^\top\mathbf{A})^{-1}\mathbf{A}^\top)\nabla(\hat{\theta}) \tag{16}$$

Using the right-hand side of equation (16) ensures that the estimating equations under the null (15) indeed hold. In practice, we observed that this adjustment leads to different degrees of modification to the gradients. We note that it may be useful and of future interest to develop a statistic based on $\mathbf{A}(\mathbf{A}^\top\mathbf{A})^{-1}\mathbf{A}^\top\nabla(\hat{\theta})$ for diagnostics of convergence.

Second, we want to account for the variance of $\hat{\theta}$ in order to develop an *efficient* score test. This affects the denominator of the score statistics in equation (13). Denote by $\hat{\theta}^{-b}$ the jackknife estimate of the parameters with the $b$-th block excluded. Recall that these values are readily available as they are used

138 to compute the jackknife estimator of the SE. Applying the jackknife variance leads to

$$Var(U_{K+1}(\theta^*)) = (B-1)Var(U_{K+1}(\hat{\theta}^{-b})), \tag{17}$$

where $B$ is the number of LD blocks or jackknife estimates, and $U_{K+1}(\hat{\theta}^{-b}) = \sum_{j \notin b} \mathbf{a}_{j,K+1} \nabla_j(\hat{\theta}^{-b})$ is the score aggregated from all except the SNPs on block $b$. However, computing $\nabla_j(\hat{\theta}^{-b})$ for every single LD block is costly and unwieldy. We propose approximating the jackknife gradient as,

$$\nabla_j(\hat{\theta}^{-b}) \approx \nabla_j(\hat{\theta}) + (\hat{\theta}^{-b} - \hat{\theta}) \cdot \left. \frac{\partial \nabla_j(\theta)}{\partial \theta} \right|_{\theta=\hat{\theta}}, \tag{18}$$

where the second term serves to explicitly account for the uncertainty in estimating $\hat{\theta}$. Importantly, both terms in equation 18 can be obtained from the null fit without involving new annotation we want to test. Now consider the derivative in the second term. Without loss of generality, consider the partial derivative with respect to the first coefficient, which is the first element of the vector $\frac{\partial \nabla_j(\theta)}{\partial \theta}$. We have

$$\frac{\partial \nabla_j(\theta)}{\partial \theta_1} = \mathbf{a}_{j,1} \left[ g''(\mathbf{a}_j^\top \theta) \ell_j' + \left( g'(\mathbf{a}_j^\top \theta) \right)^2 \ell_j'' \right]$$

where we use $\ell_j', \ell_j''$ short for $\frac{\partial \ell}{\partial g(\mathbf{a}_j^\top \theta)}$ and $\frac{\partial^2 \ell}{\partial \left( g(\mathbf{a}_j^\top \theta) \right)^2}$, respectively. This enables us to compute a jackknife score (aggregated at the LD level) that incorporates uncertainty in the estimation of $\hat{\theta}$.

$$
\begin{aligned}
\widehat{Var}(U_{K+1}(\hat{\theta}^{-b})) &= \widehat{Var}\left(\sum_{j\notin b} \mathbf{a}_{j,K+1}\nabla_j(\hat{\theta}^{-b})\right) \\
&\approx \widehat{Var}\left(\sum_{j\notin b} \mathbf{a}_{j,K+1}\left[\nabla_j(\hat{\theta}) + (\hat{\theta}^{-b} - \hat{\theta})\cdot \left.\frac{\partial \nabla_j(\theta)}{\partial \theta}\right|_{\theta=\hat{\theta}}\right]\right) \\
&\qquad\qquad \text{(first-order approximation by equation 18)} \\
&= \frac{1}{B-2}\sum_{b=1}^{B}(\mathbf{V}_b - \overline{\mathbf{V}_b})^2, \qquad \text{(sample variance of } \mathbf{V}_b \text{ across } B \text{ blocks)} \\
\text{where } \mathbf{V}_b &= \left(\sum_{j\notin b} \mathbf{a}_{j,K+1}\left[\nabla_j(\hat{\theta}) + (\hat{\theta}^{-b} - \hat{\theta})^\top \mathbf{a}_j \underbrace{\left[g''(\mathbf{a}_j^\top\hat{\theta})\ell_j' + \left(g'(\mathbf{a}_j^\top\hat{\theta})\right)^2\ell_j''\right]}_{\text{denote by } \mathbf{H}_{jj}\text{, which is a scalar}}\right]\right) \qquad (19)
\end{aligned}
$$

where we have used the empirical variance across the jackknife scores as the plug-in variance.

To facilitate the computation of the scores, we re-write the leave-one-LD-block score in equation (19) using the block-specific scores, such that each SNP-specific score gets aggregated once.

$$
\begin{aligned}
U_{K+1}(\hat{\theta}^{-b}) &= \sum_{j\notin b}\mathbf{a}_{j,K+1}\left(\nabla_j(\hat{\theta}) + \mathbf{H}_{jj}(\hat{\theta}^{-b} - \hat{\theta})^\top \mathbf{a}_j\right) \\
&= U_{K+1} - \sum_{j\in b}\mathbf{a}_{j,K+1}\nabla_j(\hat{\theta}) + \sum_{j\notin b}\mathbf{a}_{j,K+1}\mathbf{H}_{jj}(\hat{\theta}^{-b} - \hat{\theta})^\top \mathbf{a}_j \\
&\qquad\qquad \text{(Define } U_{K+1} = \textstyle\sum_{j\in[p]}\mathbf{a}_{j,K+1}\nabla_j(\hat{\theta})\text{)} \\
&= U_{K+1} - \sum_{j\in b}\mathbf{a}_{j,K+1}\nabla_j(\hat{\theta}) + (\hat{\theta}^{-b} - \hat{\theta})^\top\left[\sum_{j\in[p]}\mathbf{a}_{j,K+1}\mathbf{H}_{jj}\mathbf{a}_j - \sum_{j\in b}\mathbf{a}_{j,K+1}\mathbf{H}_{jj}\mathbf{a}_j\right], \qquad (20)
\end{aligned}
$$

where $U_{K+1}$ is the overall score based on the original parameter estimate (*i.e.*, no jackknife).

##### ***Memory cost***

In order to perform the score test, we save the following quantities from the null fit: $\nabla_j(\hat{\theta}), \mathbf{H}_{jj}$, both of which are vectors of a length that equals to the number of markers. To perform the score test (using the variance estimator that accounts for the uncertainty in $\hat{\theta}$, we also need the annotation matrix for both the baseline annotations and the new annotation for testing.

Note that the cost of calculating the score statistics is linear in the number of markers and in the number of LD blocks, so this test procedure is fast. In terms of memory requirement, $\nabla_j(\hat{\theta})$ is a vector of the same length of as a single annotation, and $\frac{\partial \nabla_j(\theta)}{\partial \theta}\Big|_{\theta=\hat{\theta}}$ has the same size as the annotation matrix (*i.e.*, number of markers by number of baseline annotations), which is equivalent to 2 annotations. Therefore, our inference procedure does not increase the memory requirement of graphREML drastically.

#### Application of graphREML to AMM

##### *Link functions*

To explicitly model the mediated heritability by the nearest genes in a given gene set, we write per-SNP heritability as the following,

$$\sigma_j^2 = f(\theta^\top \mathbf{b}_j) \circ \left(1 + f(\gamma)^\top \mathbf{a}_j\right), \tag{21}$$

where we use $\theta$ and $\gamma$ to denote the vectors of parameters for the baseline and nearest gene annotations. We compute the first derivative of the link with respect to the baseline parameters and the AMM parameters separately as the following:

$$\frac{\partial \sigma_j^2}{\partial \theta_l} = \left(1 + f(\gamma)^\top \mathbf{a}_j\right) f^{'}(\theta^\top \mathbf{b}_j)\mathbf{b}_j^l$$
$$\frac{\partial \sigma_j^2}{\partial \gamma_k} = f(\theta^\top \mathbf{b}_j) f^{'}(\gamma)\mathbf{a}_j^k$$

##### *Comparison to the original AMM*

The original AMM work models the per-SNP heritability as the following,

$$\mathbb{E}(\beta_j^2) = \tau(0) + \tau(A)\sum_k p^{(k)}\mathbf{a}_j^{(k)} = \tau(0)\left(1 + \sum_k \frac{\tau(A)}{\tau(0)} p^{(k)}\mathbf{a}_j^{(k)}\right) = \tau(0)\left(1 + \sum_k \frac{\tau^{(k)}}{\tau(0)}\mathbf{a}_j^{(k)}\right)$$

To estimate $p_k$, one first obtains estimates of $\tau^{(k)}$; then we estimate $p_k$ as $\frac{\tau^{(k)}}{\sum_l \tau^{(l)}}$.

With the new links defined in the application of graphREML to AMM, we have,

$$\mathbb{E}(\beta_j^2) = f(\boldsymbol{\theta}^\top \mathbf{b}_j)\left(1 + \sum_k f(\gamma_k)\mathbf{a}_j^{(k)}\right).$$

Analogously, we first obtain estimates of $\gamma_k$; then estimate $p_k$ with the link applied as $\frac{f(\gamma_k)}{\sum_k f(\gamma_k)}$. The key differences between the two models are that 1) graphREML enables the baseline per-SNP heritability to be variant-specific; 2) graphREML incorporates a non-negative mapping $f(\cdot)$ to ensure the non-negativity of the heritability and to enable a non-linear relationship with the annotations.
